## Supplementary material for "Accuracy of rapid point-of-care antigen-based diagnostics for SARS-CoV-2: an updated systematic review and meta-analysis with meta regression analyzing influencing factors": S1 Fig

### S1 Fig. Forest plots of all Ag-RDTs.

Caption: TP = true positive; FP = false positive; FN = false negative; TN = true negative; CI = confidence interval

**ActiveXpress**

| Author, Study ID | Sample size | TP | FN | TN | FP | Sensitivity [95%CI] | Specificity [95%CI] |
|---|---|---|---|---|---|---|---|
| FINDdx, f176.1 | 120 | 33 | 21 | 66 | 0 | 0.61 [0.47, 0.74] | 1.00 [0.95, 1.00] |

**Genedia**

| Author, Study ID | Sample size | TP | FN | TN | FP | Sensitivity [95%CI] | Specificity [95%CI] |
|---|---|---|---|---|---|---|---|
| FINDdx, f177.2 | 108 | 39 | 15 | 53 | 1 | 0.72 [0.58, 0.84] | 0.98 [0.90, 1.00] |
| FINDdx, f177.1 | 391 | 50 | 42 | 296 | 3 | 0.54 [0.44, 0.65] | 0.99 [0.97, 1.00] |

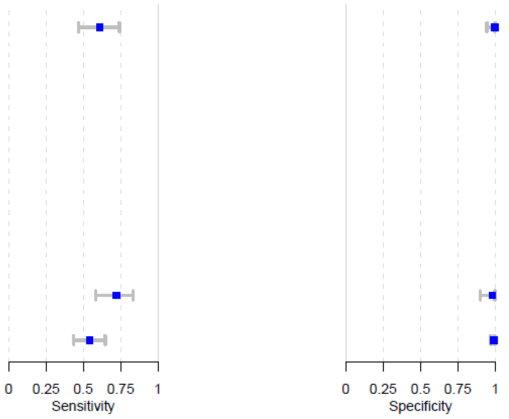


**BD Veritor**

| Author, Study ID | Sample size | TP | FN | TN | FP | Sensitivity [95%CI] | Specificity [95%CI] |
|---|---|---|---|---|---|---|---|
| Pekosz, a28.1 | 251 | 27 | 1 | 220 | 3 | 0.96 [0.82, 1.00] | 0.99 [0.96, 1.00] |
| Young, a43.1 | 251 | 29 | 9 | 212 | 1 | 0.76 [0.60, 0.89] | 1.00 [0.97, 1.00] |
| Caruana, f34.4 | 532 | 47 | 67 | 417 | 1 | 0.41 [0.32, 0.51] | 1.00 [0.99, 1.00] |
| Schuit, f64.1 | 2678 | 149 | 84 | 2436 | 9 | 0.64 [0.57, 0.70] | 1.00 [0.99, 1.00] |
| Kilic, f71.1 | 1384 | 77 | 39 | 1253 | 15 | 0.66 [0.57, 0.75] | 0.99 [0.98, 0.99] |
| Christensen, f109.1 | 278 | 52 | 8 | 217 | 1 | 0.87 [0.75, 0.94] | 1.00 [0.98, 1.00] |
| Karon, f148.1 | 347 | 131 | 66 | 150 | 0 | 0.66 [0.59, 0.73] | 1.00 [0.98, 1.00] |

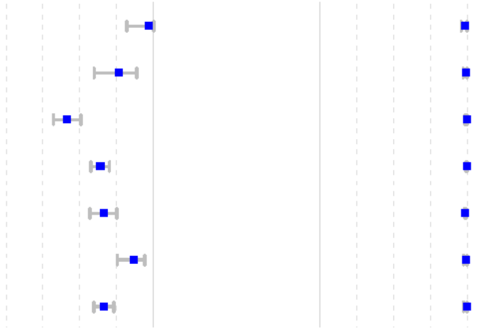

**BinaxNow**

| Author, Study ID | Sample size | TP | FN | TN | FP | Sensitivity [95%CI] | Specificity [95%CI] |
|---|---|---|---|---|---|---|---|
| Pilarowski, a29.1 | 878 | 15 | 11 | 852 | 0 | 0.58 [0.37, 0.77] | 1.00 [1.00, 1.00] |
| Pollock, f17.1 | 2308 | 226 | 66 | 2004 | 12 | 0.77 [0.72, 0.82] | 0.99 [0.99, 1.00] |
| James, f23.1 | 2339 | 86 | 66 | 2184 | 3 | 0.57 [0.48, 0.65] | 1.00 [1.00, 1.00] |
| Okoye, f51.1 | 2638 | 24 | 21 | 2593 | 0 | 0.53 [0.38, 0.68] | 1.00 [1.00, 1.00] |
| Shaikh, f105.1 | 199 | 33 | 6 | 146 | 14 | 0.85 [0.70, 0.94] | 0.91 [0.86, 0.95] |
| Shah, f133.1 | 2110 | 272 | 62 | 1769 | 7 | 0.81 [0.77, 0.86] | 1.00 [0.99, 1.00] |
| Frediani, f139.5 | 297 | 57 | 20 | 218 | 2 | 0.74 [0.63, 0.83] | 0.99 [0.97, 1.00] |
| Bachmann, f154.3 | 169 | 89 | 20 | 60 | 0 | 0.82 [0.73, 0.88] | 1.00 [0.94, 1.00] |
| Bachmann, f154.6 | 169 | 84 | 8 | 72 | 5 | 0.91 [0.84, 0.96] | 0.94 [0.86, 0.98] |
| Tinker, f156.1 | 1540 | 8 | 32 | 1500 | 0 | 0.20 [0.09, 0.36] | 1.00 [1.00, 1.00] |
| Allan−Blitz, f161.1 | 18457 | 1550 | 1603 | 15115 | 189 | 0.49 [0.47, 0.51] | 0.99 [0.99, 0.99] |

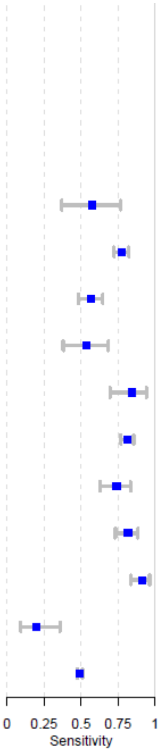


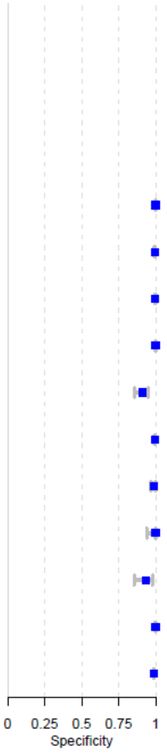


**Bioeasy**

| Author, Study ID | Sample size | TP | FN | TN | FP | Sensitivity [95%CI] | Specificity [95%CI] |
|---|---|---|---|---|---|---|---|
| Porte, a31.1 | 127 | 77 | 5 | 45 | 0 | 0.94 [0.86, 0.98] | 1.00 [0.92, 1.00] |
| Weitzel, a41.4 | 111 | 68 | 12 | 31 | 0 | 0.85 [0.75, 0.92] | 1.00 [0.89, 1.00] |
| Parada−Ricart, a58.1 | 172 | 19 | 7 | 125 | 21 | 0.73 [0.52, 0.88] | 0.86 [0.79, 0.91] |

**Rapigen**

| Author, Study ID | Sample size | TP | FN | TN | FP | Sensitivity [95%CI] | Specificity [95%CI] |
|---|---|---|---|---|---|---|---|
| Schildgen, a33.1 | 73 | 14 | 28 | 27 | 4 | 0.33 [0.20, 0.50] | 0.87 [0.70, 0.96] |
| Shrestha, a36.1 | 113 | 40 | 7 | 66 | 0 | 0.85 [0.72, 0.94] | 1.00 [0.95, 1.00] |
| Weitzel, a41.1 | 109 | 49 | 30 | 30 | 0 | 0.62 [0.50, 0.73] | 1.00 [0.88, 1.00] |
| FINDdx, a62.1 | 476 | 87 | 30 | 355 | 4 | 0.74 [0.66, 0.82] | 0.99 [0.97, 1.00] |
| FINDdx, a62.2 | 1239 | 13 | 12 | 1214 | 0 | 0.52 [0.31, 0.72] | 1.00 [1.00, 1.00] |
| Shidlovskaya, f61.1 | 106 | 44 | 34 | 28 | 0 | 0.56 [0.45, 0.68] | 1.00 [0.88, 1.00] |

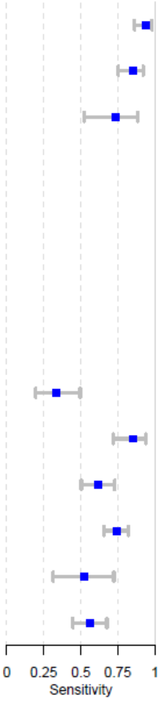


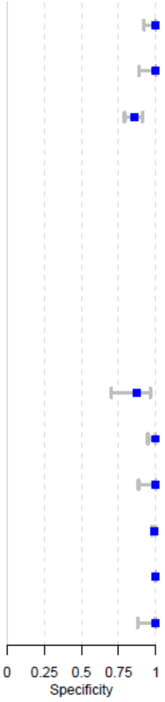

**Bio−Rad**

| Author, Study ID | Sample size | TP | FN | TN | FP | Sensitivity [95%CI] | Specificity [95%CI] |
|---|---|---|---|---|---|---|---|
| Blairon, f113.2 | 199 | 90 | 60 | 49 | 0 | 0.60 [0.52, 0.68] | 1.00 [0.93, 1.00] |

**NovaGen**

| Author, Study ID | Sample size | TP | FN | TN | FP | Sensitivity [95%CI] | Specificity [95%CI] |
|---|---|---|---|---|---|---|---|
| Blairon, f113.1 | 199 | 89 | 61 | 42 | 7 | 0.59 [0.51, 0.67] | 0.86 [0.73, 0.94] |

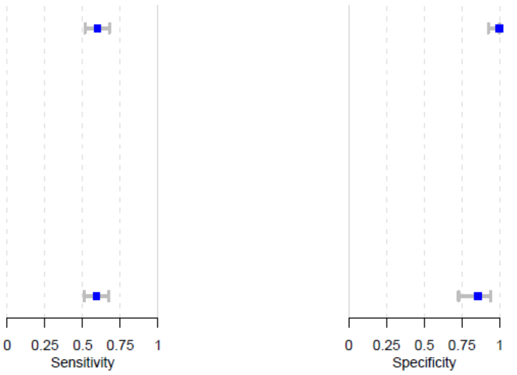


**Biotical**

| Author, Study ID | Sample size | TP | FN | TN | FP | Sensitivity [95%CI] | Specificity [95%CI] |
|---|---|---|---|---|---|---|---|
| Favresse, f31.1 | 188 | 64 | 32 | 91 | 1 | 0.67 [0.56, 0.76] | 0.99 [0.94, 1.00] |
| Van Honacker, f143.2 | 98 | 39 | 19 | 40 | 0 | 0.67 [0.54, 0.79] | 1.00 [0.91, 1.00] |

**Healgen**

| Author, Study ID | Sample size | TP | FN | TN | FP | Sensitivity [95%CI] | Specificity [95%CI] |
|---|---|---|---|---|---|---|---|
| Favresse, f31.3 | 188 | 74 | 22 | 89 | 3 | 0.77 [0.67, 0.85] | 0.97 [0.91, 0.99] |
| Seynaeve, f137.1 | 100 | 44 | 6 | 50 | 0 | 0.88 [0.76, 0.96] | 1.00 [0.93, 1.00] |

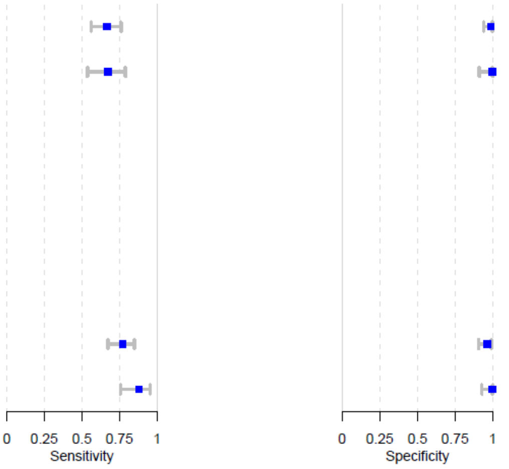


**CerTest**

| Author, Study ID | Sample size | TP | FN | TN | FP | Sensitivity [95%CI] | Specificity [95%CI] |
|---|---|---|---|---|---|---|---|
| Pérez−García, f52.1 | 320 | 91 | 79 | 150 | 0 | 0.54 [0.46, 0.61] | 1.00 [0.98, 1.00] |
| Koeleman, f103.3 | 80 | 22 | 18 | 39 | 1 | 0.55 [0.38, 0.71] | 0.98 [0.87, 1.00] |

**CareStart**

| Author, Study ID | Sample size | TP | FN | TN | FP | Sensitivity [95%CI] | Specificity [95%CI] |
|---|---|---|---|---|---|---|---|
| Pollock, f59.1 | 1498 | 135 | 99 | 1243 | 21 | 0.58 [0.51, 0.64] | 0.98 [0.98, 0.99] |

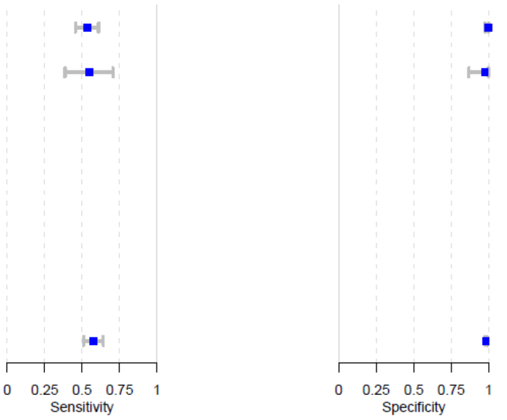

**Ecotest**

| Author, Study ID | Sample size | TP | FN | TN | FP | Sensitivity [95%CI] | Specificity [95%CI] |
|---|---|---|---|---|---|---|---|
| Homza, f87.1 | 318 | 81 | 26 | 204 | 7 | 0.76 [0.66, 0.84] | 0.97 [0.93, 0.99] |
| Homza, f99.1 | 494 | 125 | 39 | 321 | 9 | 0.76 [0.69, 0.82] | 0.97 [0.95, 0.99] |

**ND COVID**

| Author, Study ID | Sample size | TP | FN | TN | FP | Sensitivity [95%CI] | Specificity [95%CI] |
|---|---|---|---|---|---|---|---|
| Homza, f87.3 | 191 | 54 | 23 | 64 | 50 | 0.70 [0.59, 0.80] | 0.56 [0.47, 0.65] |

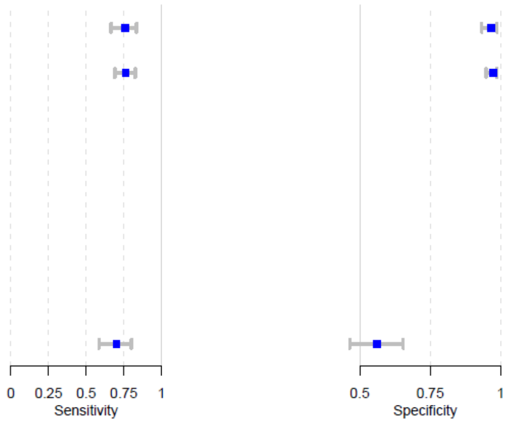


**Encode**

| Author, Study ID | Sample size | TP | FN | TN | FP | Sensitivity [95%CI] | Specificity [95%CI] |
|---|---|---|---|---|---|---|---|
| Pickering, f73.5 | 200 | 74 | 26 | 100 | 0 | 0.74 [0.64, 0.82] | 1.00 [0.96, 1.00] |

**Spring**

| Author, Study ID | Sample size | TP | FN | TN | FP | Sensitivity [95%CI] | Specificity [95%CI] |
|---|---|---|---|---|---|---|---|
| Pickering, f73.4 | 200 | 77 | 23 | 98 | 2 | 0.77 [0.68, 0.85] | 0.98 [0.93, 1.00] |

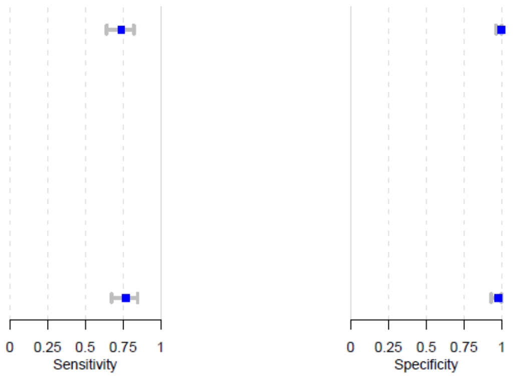

**Espline**

| Author, Study ID | Sample size | TP | FN | TN | FP | Sensitivity [95%CI] | Specificity [95%CI] |
|---|---|---|---|---|---|---|---|
| Takeda, a50.1 | 162 | 50 | 12 | 100 | 0 | 0.81 [0.69, 0.90] | 1.00 [0.96, 1.00] |
| Sberna, f83.1 | 136 | 5 | 57 | 74 | 0 | 0.08 [0.03, 0.18] | 1.00 [0.95, 1.00] |
| FINDdx, f92.1 | 723 | 88 | 24 | 611 | 0 | 0.79 [0.70, 0.86] | 1.00 [0.99, 1.00] |

**Standard Q nasal**

| Author, Study ID | Sample size | TP | FN | TN | FP | Sensitivity [95%CI] | Specificity [95%CI] |
|---|---|---|---|---|---|---|---|
| Lindner, a53.2 | 179 | 33 | 8 | 136 | 2 | 0.80 [0.65, 0.91] | 0.99 [0.95, 1.00] |
| Lindner, f15.2 | 144 | 33 | 7 | 104 | 0 | 0.82 [0.67, 0.93] | 1.00 [0.96, 1.00] |
| Nikolai, f35.1 | 132 | 31 | 5 | 96 | 0 | 0.86 [0.70, 0.95] | 1.00 [0.96, 1.00] |
| Nikolai, f35.2 | 132 | 31 | 5 | 96 | 0 | 0.86 [0.70, 0.95] | 1.00 [0.96, 1.00] |
| Nikolai, f35.4 | 96 | 31 | 3 | 61 | 1 | 0.91 [0.76, 0.98] | 0.98 [0.91, 1.00] |
| Stohr, f45.2 | 1588 | 118 | 74 | 1392 | 4 | 0.62 [0.54, 0.68] | 1.00 [0.99, 1.00] |
| Agarwal, f153.1 | 467 | 26 | 3 | 436 | 2 | 0.90 [0.73, 0.98] | 1.00 [0.98, 1.00] |
| FINDdx, f182.2 | 214 | 66 | 12 | 135 | 1 | 0.85 [0.75, 0.92] | 0.99 [0.96, 1.00] |

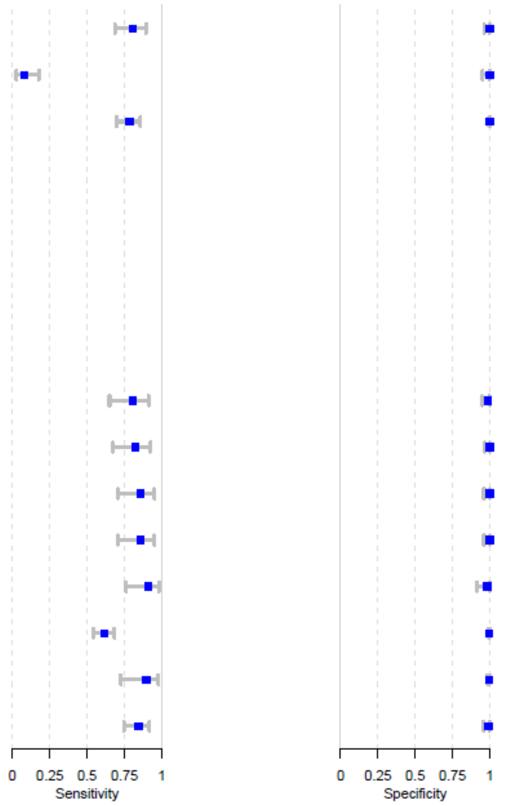


**Exdia**

| Author, Study ID | Sample size | TP | FN | TN | FP | Sensitivity [95%CI] | Specificity [95%CI] |
|---|---|---|---|---|---|---|---|
| Caruana, f34.3 | 532 | 55 | 59 | 416 | 2 | 0.48 [0.39, 0.58] | 1.00 [0.98, 1.00] |

**Ichroma**

| Author, Study ID | Sample size | TP | FN | TN | FP | Sensitivity [95%CI] | Specificity [95%CI] |
|---|---|---|---|---|---|---|---|
| FINDdx, f39.1 | 232 | 30 | 11 | 191 | 0 | 0.73 [0.57, 0.86] | 1.00 [0.98, 1.00] |

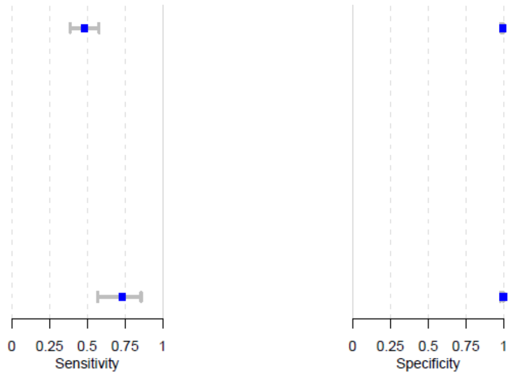

**Flowflex LF**

| Author, Study ID | Sample size | TP | FN | TN | FP | Sensitivity [95%CI] | Specificity [95%CI] |
|---|---|---|---|---|---|---|---|
| Karon, f148.4 | 347 | 153 | 44 | 146 | 4 | 0.78 [0.71, 0.83] | 0.97 [0.93, 0.99] |
| FINDdx, f175.1 | 279 | 58 | 5 | 215 | 1 | 0.92 [0.82, 0.97] | 1.00 [0.97, 1.00] |

**QuickChaser**

| Author, Study ID | Sample size | TP | FN | TN | FP | Sensitivity [95%CI] | Specificity [95%CI] |
|---|---|---|---|---|---|---|---|
| Kurihara, f152.1 | 1401 | 62 | 21 | 1316 | 2 | 0.75 [0.64, 0.84] | 1.00 [1.00, 1.00] |

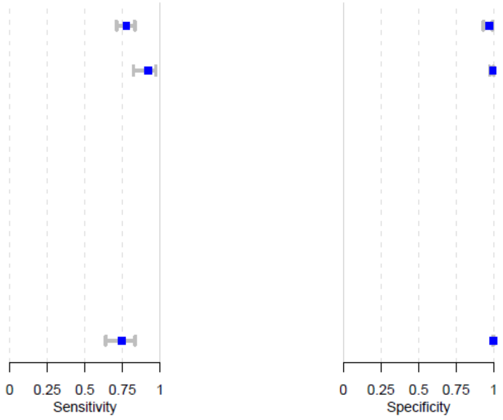


**Frend**

| Author, Study ID | Sample size | TP | FN | TN | FP | Sensitivity [95%CI] | Specificity [95%CI] |
|---|---|---|---|---|---|---|---|
| Orsi, f191.1 | 110 | 56 | 4 | 50 | 0 | 0.93 [0.84, 0.98] | 1.00 [0.93, 1.00] |

**RapidTesta**

| Author, Study ID | Sample size | TP | FN | TN | FP | Sensitivity [95%CI] | Specificity [95%CI] |
|---|---|---|---|---|---|---|---|
| Suzuki, f194.2 | 1127 | 58 | 16 | 1028 | 25 | 0.78 [0.67, 0.87] | 0.98 [0.96, 0.98] |
| Suzuki, f194.1 | 1127 | 53 | 21 | 1045 | 8 | 0.72 [0.60, 0.81] | 0.99 [0.98, 1.00] |

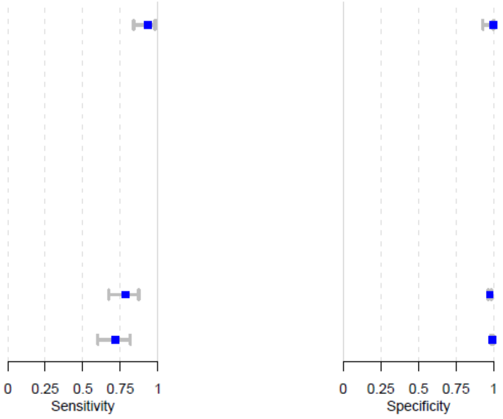


**GenBody**

| Author, Study ID | Sample size | TP | FN | TN | FP | Sensitivity [95%CI] | Specificity [95%CI] |
|---|---|---|---|---|---|---|---|
| Kim, f186.1 | 130 | 27 | 3 | 98 | 2 | 0.90 [0.74, 0.98] | 0.98 [0.93, 1.00] |
| Kim, f186.2 | 200 | 94 | 6 | 100 | 0 | 0.94 [0.87, 0.98] | 1.00 [0.96, 1.00] |

**NG Biotech**

| Author, Study ID | Sample size | TP | FN | TN | FP | Sensitivity [95%CI] | Specificity [95%CI] |
|---|---|---|---|---|---|---|---|
| Fourati, f190.5 | 634 | 97 | 200 | 332 | 5 | 0.33 [0.27, 0.38] | 0.98 [0.97, 1.00] |

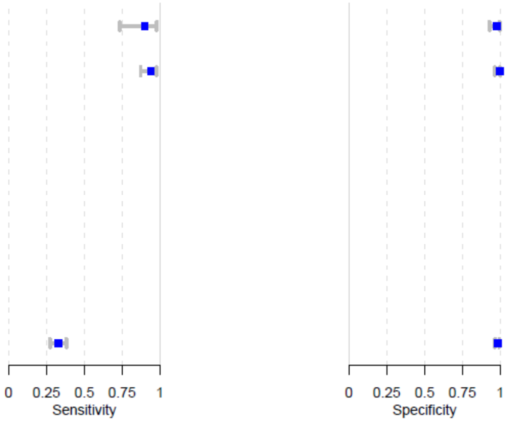

**Hotgen**

| Author, Study ID | Sample size | TP | FN | TN | FP | Sensitivity [95%CI] | Specificity [95%CI] |
|---|---|---|---|---|---|---|---|
| FINDdx, f178.1 | 454 | 94 | 12 | 348 | 0 | 0.89 [0.81, 0.94] | 1.00 [0.99, 1.00] |

**Sure Status**

| Author, Study ID | Sample size | TP | FN | TN | FP | Sensitivity [95%CI] | Specificity [95%CI] |
|---|---|---|---|---|---|---|---|
| FINDdx, f180.1 | 519 | 91 | 9 | 407 | 12 | 0.91 [0.84, 0.96] | 0.97 [0.95, 0.98] |
| FINDdx, f180.2 | 997 | 102 | 28 | 865 | 2 | 0.78 [0.70, 0.85] | 1.00 [0.99, 1.00] |

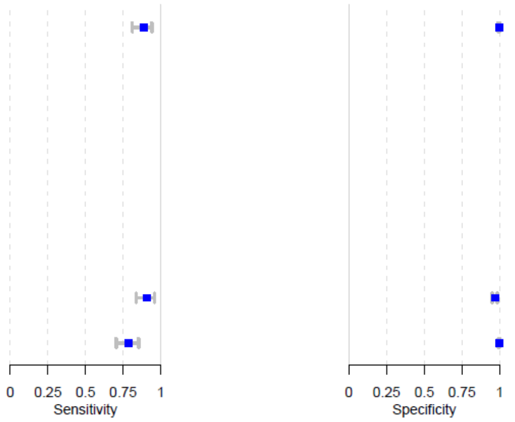


**Innova**

| Author, Study ID | Sample size | TP | FN | TN | FP | Sensitivity [95%CI] | Specificity [95%CI] |
|---|---|---|---|---|---|---|---|
| Houston, f25.1 | 728 | 242 | 38 | 426 | 22 | 0.86 [0.82, 0.90] | 0.95 [0.93, 0.97] |
| Young, f56.1 | 786 | 133 | 81 | 572 | 0 | 0.62 [0.55, 0.69] | 1.00 [0.99, 1.00] |
| Pickering, f73.1 | 200 | 89 | 11 | 99 | 1 | 0.89 [0.81, 0.94] | 0.99 [0.95, 1.00] |
| García−Fiñana, f131.1 | 5504 | 28 | 42 | 5431 | 3 | 0.40 [0.28, 0.52] | 1.00 [1.00, 1.00] |

**CLINITEST**

| Author, Study ID | Sample size | TP | FN | TN | FP | Sensitivity [95%CI] | Specificity [95%CI] |
|---|---|---|---|---|---|---|---|
| Torres, f29.1 | 178 | 73 | 18 | 87 | 0 | 0.80 [0.71, 0.88] | 1.00 [0.96, 1.00] |
| Torres, f29.2 | 92 | 15 | 10 | 67 | 0 | 0.60 [0.39, 0.79] | 1.00 [0.95, 1.00] |
| Baro, f33.2 | 286 | 52 | 49 | 182 | 3 | 0.52 [0.41, 0.62] | 0.98 [0.95, 1.00] |
| Merino−Amador, f160.1 | 450 | 179 | 13 | 256 | 2 | 0.93 [0.89, 0.96] | 0.99 [0.97, 1.00] |

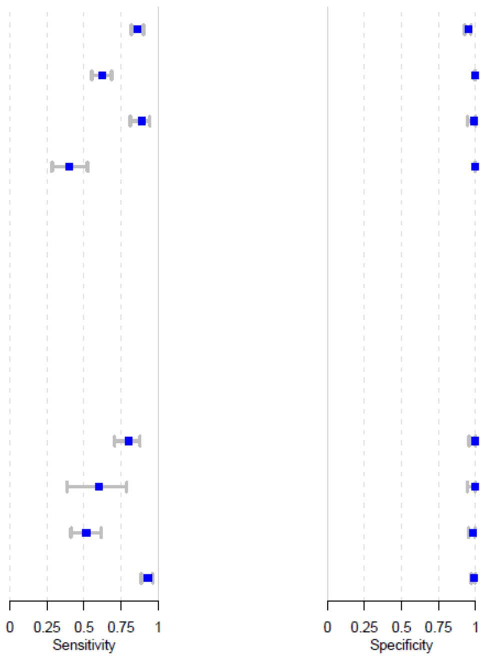

**Joysbio**

| Author, Study ID | Sample size | TP | FN | TN | FP | Sensitivity [95%CI] | Specificity [95%CI] |
|---|---|---|---|---|---|---|---|
| FINDdx, f40.1 | 265 | 31 | 13 | 219 | 2 | 0.70 [0.55, 0.83] | 0.99 [0.97, 1.00] |
| Homza, f87.2 | 225 | 52 | 38 | 133 | 2 | 0.58 [0.47, 0.68] | 0.98 [0.95, 1.00] |

**Wondfo**

| Author, Study ID | Sample size | TP | FN | TN | FP | Sensitivity [95%CI] | Specificity [95%CI] |
|---|---|---|---|---|---|---|---|
| FINDdx, f41.1 | 328 | 48 | 8 | 272 | 0 | 0.86 [0.74, 0.94] | 1.00 [0.99, 1.00] |

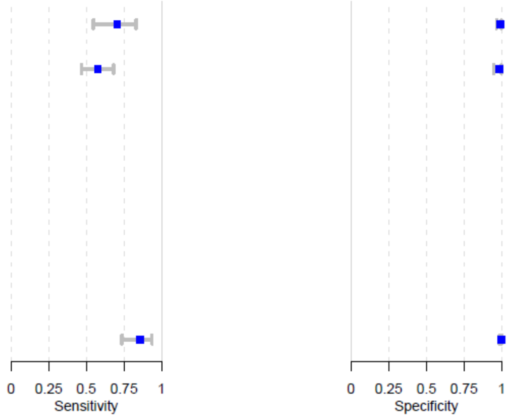


**Lituo**

| Author, Study ID | Sample size | TP | FN | TN | FP | Sensitivity [95%CI] | Specificity [95%CI] |
|---|---|---|---|---|---|---|---|
| Terpos, f130.1 | 358 | 102 | 12 | 243 | 1 | 0.90 [0.82, 0.94] | 1.00 [0.98, 1.00] |

**Fluorecare**

| Author, Study ID | Sample size | TP | FN | TN | FP | Sensitivity [95%CI] | Specificity [95%CI] |
|---|---|---|---|---|---|---|---|
| Lunca, f138.1 | 47 | 23 | 17 | 7 | 0 | 0.58 [0.41, 0.73] | 1.00 [0.59, 1.00] |

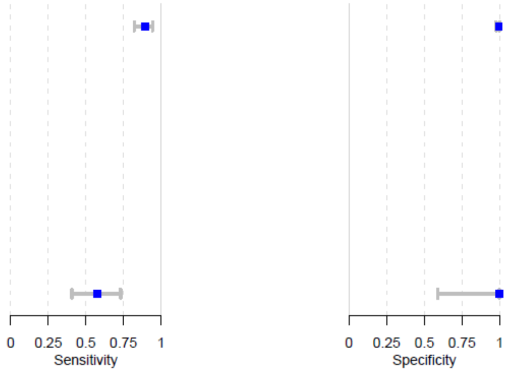


**LumiraDx**

| Author, Study ID | Sample size | TP | FN | TN | FP | Sensitivity [95%CI] | Specificity [95%CI] |
|---|---|---|---|---|---|---|---|
| Kohmer, f32.4 | 100 | 37 | 37 | 26 | 0 | 0.50 [0.38, 0.62] | 1.00 [0.87, 1.00] |
| Drain, f43.1 | 257 | 81 | 2 | 168 | 6 | 0.98 [0.92, 1.00] | 0.97 [0.93, 0.99] |
| Drain, f43.2 | 255 | 39 | 1 | 210 | 5 | 0.98 [0.87, 1.00] | 0.98 [0.95, 0.99] |
| Krüger, f58.1 | 761 | 120 | 26 | 611 | 4 | 0.82 [0.75, 0.88] | 0.99 [0.98, 1.00] |
| Bianco, f102.1 | 907 | 269 | 29 | 561 | 48 | 0.90 [0.86, 0.93] | 0.92 [0.90, 0.94] |
| Caramello, f108.2 | 149 | 84 | 13 | 46 | 6 | 0.87 [0.78, 0.93] | 0.88 [0.77, 0.96] |
| Blairon, f113.3 | 198 | 91 | 58 | 49 | 0 | 0.61 [0.53, 0.69] | 1.00 [0.93, 1.00] |
| Fernández, f128.1 | 46 | 21 | 3 | 22 | 0 | 0.88 [0.68, 0.97] | 1.00 [0.85, 1.00] |
| Karon, f148.2 | 347 | 164 | 33 | 150 | 0 | 0.83 [0.77, 0.88] | 1.00 [0.98, 1.00] |
| Leli, f151.1 | 792 | 114 | 52 | 596 | 30 | 0.69 [0.61, 0.76] | 0.95 [0.93, 0.97] |
| Cento, f196.1 | 960 | 297 | 50 | 596 | 17 | 0.86 [0.81, 0.89] | 0.97 [0.96, 0.98] |

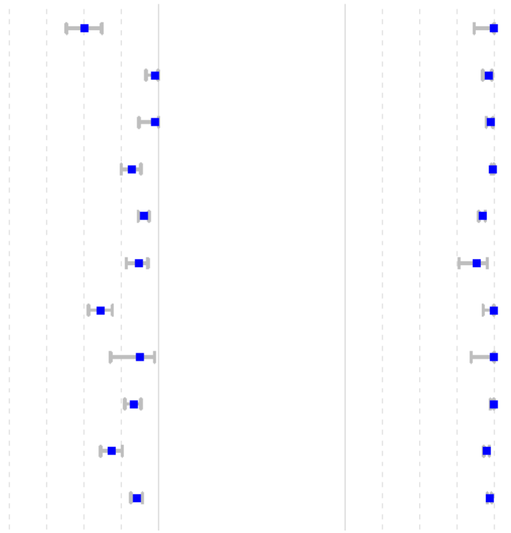

**Nadal**

| Author, Study ID | Sample size | TP | FN | TN | FP | Sensitivity [95%CI] | Specificity [95%CI] |
|---|---|---|---|---|---|---|---|
| Kohmer, f32.3 | 100 | 18 | 56 | 26 | 0 | 0.24 [0.15, 0.36] | 1.00 [0.87, 1.00] |
| FINDdx, f94.1 | 462 | 61 | 8 | 390 | 3 | 0.88 [0.78, 0.95] | 0.99 [0.98, 1.00] |
| Ifko, f169.1 | 125 | 20 | 3 | 90 | 12 | 0.87 [0.66, 0.97] | 0.88 [0.80, 0.94] |

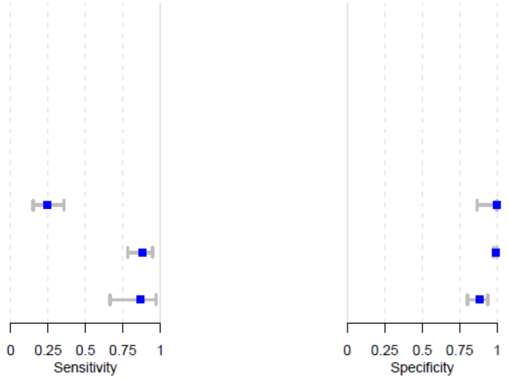


**NowCheck nasal**

| Author, Study ID | Sample size | TP | FN | TN | FP | Sensitivity [95%CI] | Specificity [95%CI] |
|---|---|---|---|---|---|---|---|
| FINDdx, f91.1 | 218 | 71 | 8 | 137 | 2 | 0.90 [0.81, 0.96] | 0.99 [0.95, 1.00] |

**Mologic**

| Author, Study ID | Sample size | TP | FN | TN | FP | Sensitivity [95%CI] | Specificity [95%CI] |
|---|---|---|---|---|---|---|---|
| FINDdx, f93.1 | 665 | 176 | 18 | 471 | 0 | 0.91 [0.86, 0.94] | 1.00 [0.99, 1.00] |

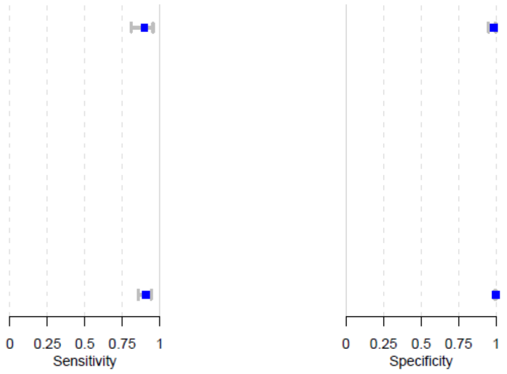


**OA−LFA**

| Author, Study ID | Sample size | TP | FN | TN | FP | Sensitivity [95%CI] | Specificity [95%CI] |
|---|---|---|---|---|---|---|---|
| Bachmann, f154.1 | 167 | 75 | 33 | 57 | 2 | 0.69 [0.60, 0.78] | 0.97 [0.88, 1.00] |
| Bachmann, f154.4 | 167 | 75 | 15 | 75 | 2 | 0.83 [0.74, 0.90] | 0.97 [0.91, 1.00] |

**Afias**

| Author, Study ID | Sample size | TP | FN | TN | FP | Sensitivity [95%CI] | Specificity [95%CI] |
|---|---|---|---|---|---|---|---|
| Baccani, f165.3 | 81 | 9 | 15 | 57 | 0 | 0.38 [0.19, 0.59] | 1.00 [0.94, 1.00] |

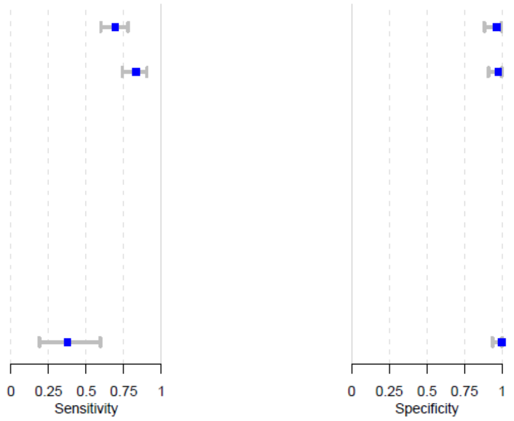

**Orient Gene**

| Author, Study ID | Sample size | TP | FN | TN | FP | Sensitivity [95%CI] | Specificity [95%CI] |
|---|---|---|---|---|---|---|---|
| Nordgren, f117.2 | 332 | 124 | 32 | 131 | 45 | 0.80 [0.72, 0.86] | 0.74 [0.67, 0.81] |
| Van Honacker, f143.3 | 98 | 48 | 10 | 37 | 3 | 0.83 [0.71, 0.91] | 0.92 [0.80, 0.98] |

**AMP**

| Author, Study ID | Sample size | TP | FN | TN | FP | Sensitivity [95%CI] | Specificity [95%CI] |
|---|---|---|---|---|---|---|---|
| Leixner, f125.1 | 392 | 65 | 29 | 297 | 1 | 0.69 [0.59, 0.78] | 1.00 [0.98, 1.00] |

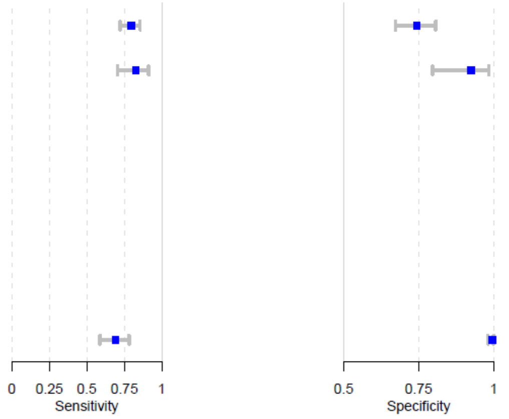


**Panbio (nasal)**

| Author, Study ID | Sample size | TP | FN | TN | FP | Sensitivity [95%CI] | Specificity [95%CI] |
|---|---|---|---|---|---|---|---|
| FINDdx, f42.1 | 281 | 38 | 6 | 235 | 2 | 0.86 [0.73, 0.95] | 0.99 [0.97, 1.00] |

**BD Veritor nasal**

| Author, Study ID | Sample size | TP | FN | TN | FP | Sensitivity [95%CI] | Specificity [95%CI] |
|---|---|---|---|---|---|---|---|
| Stohr, f45.1 | 1565 | 86 | 90 | 1387 | 2 | 0.49 [0.41, 0.56] | 1.00 [1.00, 1.00] |

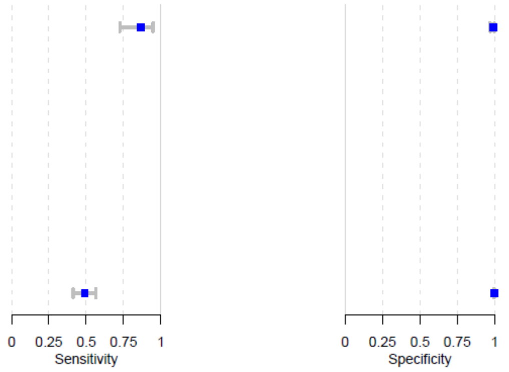


**QuickNavi**

| Author, Study ID | Sample size | TP | FN | TN | FP | Sensitivity [95%CI] | Specificity [95%CI] |
|---|---|---|---|---|---|---|---|
| Takeuchi, f12.1 | 1186 | 91 | 14 | 1081 | 0 | 0.87 [0.79, 0.92] | 1.00 [1.00, 1.00] |
| Takeuchi, f60.1 | 862 | 37 | 14 | 811 | 0 | 0.72 [0.58, 0.84] | 1.00 [1.00, 1.00] |

**ECODiagnostica**

| Author, Study ID | Sample size | TP | FN | TN | FP | Sensitivity [95%CI] | Specificity [95%CI] |
|---|---|---|---|---|---|---|---|
| Filgueiras, f14.1 | 139 | 38 | 17 | 83 | 1 | 0.69 [0.55, 0.81] | 0.99 [0.94, 1.00] |

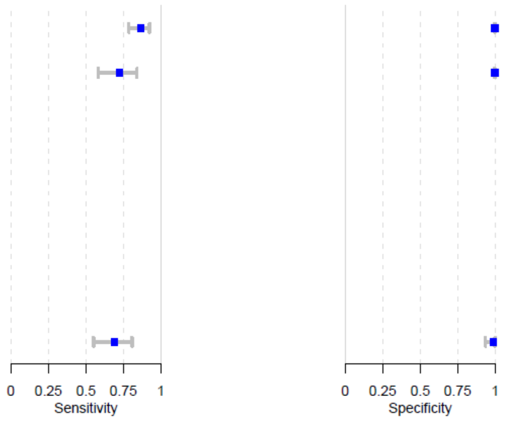

**R−Biopharm**

| Author, Study ID | Sample size | TP | FN | TN | FP | Sensitivity [95%CI] | Specificity [95%CI] |
|---|---|---|---|---|---|---|---|
| Toptan, a55.2 | 70 | 16 | 16 | 38 | 0 | 0.50 [0.32, 0.68] | 1.00 [0.91, 1.00] |
| Toptan, a55.1 | 67 | 45 | 13 | 9 | 0 | 0.78 [0.65, 0.88] | 1.00 [0.66, 1.00] |
| Kohmer, f32.1 | 100 | 29 | 45 | 25 | 1 | 0.39 [0.28, 0.51] | 0.96 [0.80, 1.00] |

**NowCheck**

| Author, Study ID | Sample size | TP | FN | TN | FP | Sensitivity [95%CI] | Specificity [95%CI] |
|---|---|---|---|---|---|---|---|
| FINDdx, a61.1 | 400 | 91 | 11 | 290 | 8 | 0.89 [0.81, 0.94] | 0.97 [0.95, 0.99] |
| FINDdx, f91.2 | 218 | 71 | 8 | 137 | 2 | 0.90 [0.81, 0.96] | 0.99 [0.95, 1.00] |
| Onsongo, f174.1 | 997 | 109 | 43 | 824 | 21 | 0.72 [0.64, 0.79] | 0.98 [0.96, 0.98] |

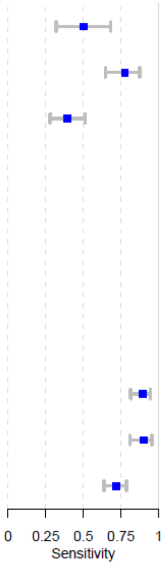


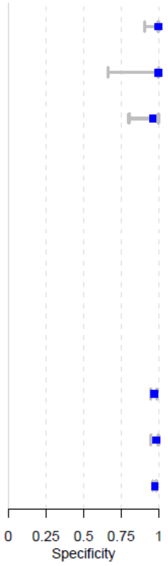


**Romed**

| Author, Study ID | Sample size | TP | FN | TN | FP | Sensitivity [95%CI] | Specificity [95%CI] |
|---|---|---|---|---|---|---|---|
| Koeleman, f103.6 | 900 | 220 | 80 | 599 | 1 | 0.73 [0.68, 0.78] | 1.00 [0.99, 1.00] |
| Koeleman, f103.1 | 80 | 29 | 11 | 40 | 0 | 0.72 [0.56, 0.85] | 1.00 [0.91, 1.00] |

**Indicaid**

| Author, Study ID | Sample size | TP | FN | TN | FP | Sensitivity [95%CI] | Specificity [95%CI] |
|---|---|---|---|---|---|---|---|
| Chiu, f107.1 | 349 | 64 | 11 | 260 | 14 | 0.85 [0.75, 0.92] | 0.95 [0.92, 0.97] |
| Chiu, f107.2 | 349 | 62 | 13 | 264 | 10 | 0.83 [0.72, 0.90] | 0.96 [0.93, 0.98] |
| Chiu, f107.3 | 22994 | 32 | 6 | 22938 | 18 | 0.84 [0.69, 0.94] | 1.00 [1.00, 1.00] |

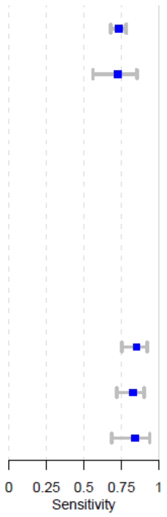


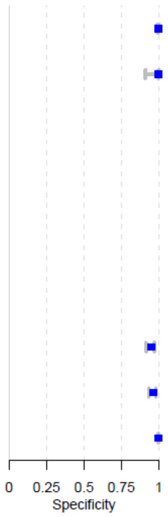

**SGTI−flex**

| Author, Study ID | Sample size | TP | FN | TN | FP | Sensitivity [95%CI] | Specificity [95%CI] |
|---|---|---|---|---|---|---|---|
| Shidlovskaya, f61.2 | 106 | 41 | 37 | 27 | 1 | 0.53 [0.41, 0.64] | 0.96 [0.82, 1.00] |

**Sienna**

| Author, Study ID | Sample size | TP | FN | TN | FP | Sensitivity [95%CI] | Specificity [95%CI] |
|---|---|---|---|---|---|---|---|
| Bouassa, f67.1 | 150 | 90 | 10 | 50 | 0 | 0.90 [0.82, 0.95] | 1.00 [0.93, 1.00] |

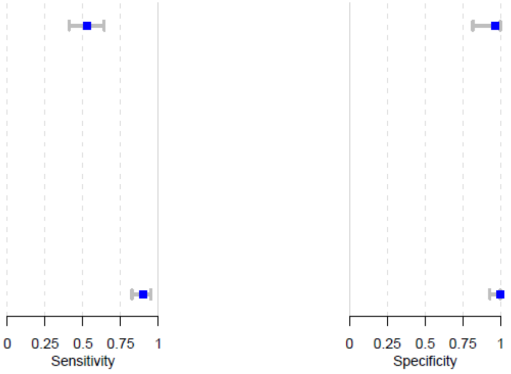


**Sofia**

| Author, Study ID | Sample size | TP | FN | TN | FP | Sensitivity [95%CI] | Specificity [95%CI] |
|---|---|---|---|---|---|---|---|
| Beck, a04.1 | 346 | 47 | 14 | 284 | 1 | 0.77 [0.64, 0.87] | 1.00 [0.98, 1.00] |
| Porte, a32.1 | 64 | 30 | 2 | 31 | 1 | 0.94 [0.79, 0.99] | 0.97 [0.84, 1.00] |
| Herrera, a46.1 | 1172 | 352 | 107 | 707 | 6 | 0.77 [0.72, 0.80] | 0.99 [0.98, 1.00] |
| Marti, f46.1 | 427 | 31 | 12 | 383 | 1 | 0.72 [0.56, 0.85] | 1.00 [0.99, 1.00] |
| Jääskeläinen, f50.1 | 188 | 119 | 29 | 40 | 0 | 0.80 [0.73, 0.86] | 1.00 [0.91, 1.00] |
| Brihn, f98.1 | 2039 | 98 | 51 | 1878 | 12 | 0.66 [0.58, 0.73] | 0.99 [0.99, 1.00] |
| Bornemann, f112.1 | 1391 | 52 | 39 | 1291 | 9 | 0.57 [0.46, 0.68] | 0.99 [0.99, 1.00] |
| Harris, f114.1 | 885 | 251 | 54 | 573 | 7 | 0.82 [0.78, 0.86] | 0.99 [0.98, 1.00] |
| Smith, f122.1 | 2887 | 180 | 55 | 2645 | 7 | 0.77 [0.71, 0.82] | 1.00 [1.00, 1.00] |
| Bachmann, f154.2 | 170 | 81 | 29 | 59 | 1 | 0.74 [0.64, 0.82] | 0.98 [0.91, 1.00] |
| Bachmann, f154.5 | 170 | 79 | 13 | 75 | 3 | 0.86 [0.77, 0.92] | 0.96 [0.89, 0.99] |

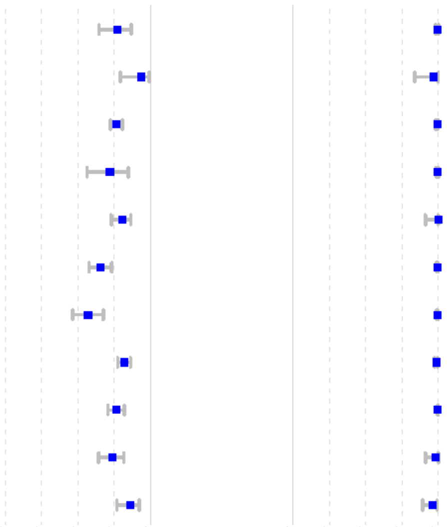

**Standard Q**

| Author, Study ID | Sample size | TP | FN | TN | FP | Sensitivity [95%CI] | Specificity [95%CI] |
|---|---|---|---|---|---|---|---|
| Berger, a05.2 | 529 | 170 | 21 | 337 | 1 | 0.89 [0.84, 0.93] | 1.00 [0.98, 1.00] |
| Cerutti, a08.1 | 185 | 75 | 29 | 81 | 0 | 0.72 [0.62, 0.80] | 1.00 [0.96, 1.00] |
| Gupta, a13.1 | 330 | 63 | 14 | 252 | 1 | 0.82 [0.71, 0.90] | 1.00 [0.98, 1.00] |
| Igl...i, a15.1 | 970 | 158 | 28 | 780 | 4 | 0.85 [0.79, 0.90] | 1.00 [0.99, 1.00] |
| Krüttgen, a16.1 | 150 | 53 | 22 | 72 | 3 | 0.71 [0.59, 0.81] | 0.96 [0.89, 0.99] |
| Krüger, a17.1 | 1263 | 36 | 11 | 1207 | 9 | 0.77 [0.62, 0.88] | 0.99 [0.99, 1.00] |
| Lindner, a21.1 | 289 | 29 | 10 | 248 | 2 | 0.74 [0.58, 0.87] | 0.99 [0.97, 1.00] |

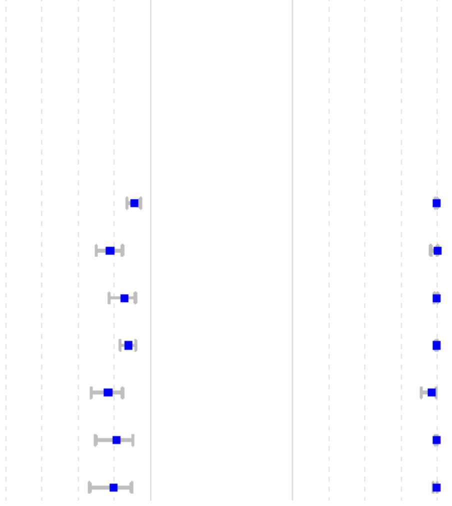

| | | | | | | | |
|---|---|---|---|---|---|---|---|
| Lindner, a21.2 | 289 | 31 | 8 | 249 | 1 | 0.80 [0.64, 0.91] | 1.00 [0.98, 1.00] |
| Nalumansi, a27.1 | 262 | 63 | 27 | 159 | 13 | 0.70 [0.59, 0.79] | 0.92 [0.87, 0.96] |
| Schildgen, a33.3 | 73 | 37 | 5 | 6 | 25 | 0.88 [0.74, 0.96] | 0.19 [0.07, 0.38] |
| Schwob, a35.1 | 333 | 104 | 8 | 221 | 0 | 0.93 [0.86, 0.97] | 1.00 [0.98, 1.00] |
| Lindner, a53.1 | 179 | 30 | 11 | 137 | 1 | 0.73 [0.57, 0.86] | 0.99 [0.96, 1.00] |
| Chaimao, a57.1 | 454 | 59 | 1 | 389 | 5 | 0.98 [0.91, 1.00] | 0.99 [0.97, 1.00] |
| FINDdx, a64.1 | 400 | 94 | 12 | 287 | 7 | 0.89 [0.81, 0.94] | 0.98 [0.95, 0.99] |
| Turcato, f09.1 | 3410 | 179 | 44 | 3157 | 30 | 0.80 [0.74, 0.85] | 0.99 [0.99, 0.99] |
| Lindner, f15.1 | 146 | 34 | 6 | 105 | 1 | 0.85 [0.70, 0.94] | 0.99 [0.95, 1.00] |
| Möckel, f19.1 | 271 | 67 | 22 | 182 | 0 | 0.75 [0.65, 0.84] | 1.00 [0.98, 1.00] |
| Möckel, f19.2 | 202 | 18 | 7 | 176 | 1 | 0.72 [0.51, 0.88] | 0.99 [0.97, 1.00] |
| Osterman, f20.2 | 642 | 165 | 91 | 377 | 9 | 0.64 [0.58, 0.70] | 0.98 [0.96, 0.99] |
| Kannian, f26.1 | 37 | 15 | 12 | 10 | 0 | 0.56 [0.35, 0.74] | 1.00 [0.69, 1.00] |
| Favresse, f31.4 | 188 | 67 | 29 | 92 | 0 | 0.70 [0.60, 0.79] | 1.00 [0.96, 1.00] |
| Kohmer, f32.2 | 100 | 32 | 42 | 26 | 0 | 0.43 [0.32, 0.55] | 1.00 [0.87, 1.00] |
| Baro, f33.3 | 286 | 44 | 57 | 178 | 7 | 0.44 [0.34, 0.54] | 0.96 [0.92, 0.98] |
| Caruana, f34.1 | 532 | 47 | 67 | 417 | 1 | 0.41 [0.32, 0.51] | 1.00 [0.99, 1.00] |
| Nikolai, f35.3 | 96 | 31 | 3 | 62 | 0 | 0.91 [0.76, 0.98] | 1.00 [0.94, 1.00] |
| Pena, f36.1 | 842 | 51 | 22 | 766 | 3 | 0.70 [0.58, 0.80] | 1.00 [0.99, 1.00] |
| Ristic, f44.1 | 120 | 25 | 18 | 77 | 0 | 0.58 [0.42, 0.73] | 1.00 [0.95, 1.00] |
| Jääskeläinen, f50.2 | 198 | 128 | 30 | 40 | 0 | 0.81 [0.74, 0.87] | 1.00 [0.91, 1.00] |
| Salvagno, f54.1 | 321 | 108 | 41 | 171 | 1 | 0.72 [0.65, 0.80] | 0.99 [0.97, 1.00] |
| Schuit, f64.2 | 1596 | 83 | 49 | 1456 | 8 | 0.63 [0.54, 0.71] | 1.00 [0.99, 1.00] |
| Kernéis, f69.1 | 1109 | 81 | 5 | 1013 | 10 | 0.94 [0.87, 0.98] | 0.99 [0.98, 1.00] |
| Thell, f81.1 | 541 | 171 | 42 | 325 | 3 | 0.80 [0.74, 0.85] | 0.99 [0.97, 1.00] |
| Homza, f87.4 | 139 | 26 | 16 | 96 | 1 | 0.62 [0.46, 0.76] | 0.99 [0.94, 1.00] |
| Holzner, f97.1 | 2375 | 379 | 172 | 1816 | 8 | 0.69 [0.65, 0.73] | 1.00 [0.99, 1.00] |
| Koeleman, f103.2 | 80 | 25 | 15 | 35 | 5 | 0.62 [0.46, 0.77] | 0.88 [0.73, 0.96] |
| Caramello, f108.1 | 175 | 84 | 29 | 61 | 1 | 0.74 [0.65, 0.82] | 0.98 [0.91, 1.00] |
| Abdul−Mumin, f123.1 | 193 | 27 | 15 | 123 | 28 | 0.64 [0.48, 0.78] | 0.81 [0.74, 0.87] |
| Fernandez−Montero, f126.1 | 2543 | 35 | 14 | 2486 | 8 | 0.71 [0.57, 0.83] | 1.00 [0.99, 1.00] |
| Lee, f136.1 | 680 | 109 | 271 | 300 | 0 | 0.29 [0.24, 0.34] | 1.00 [0.99, 1.00] |

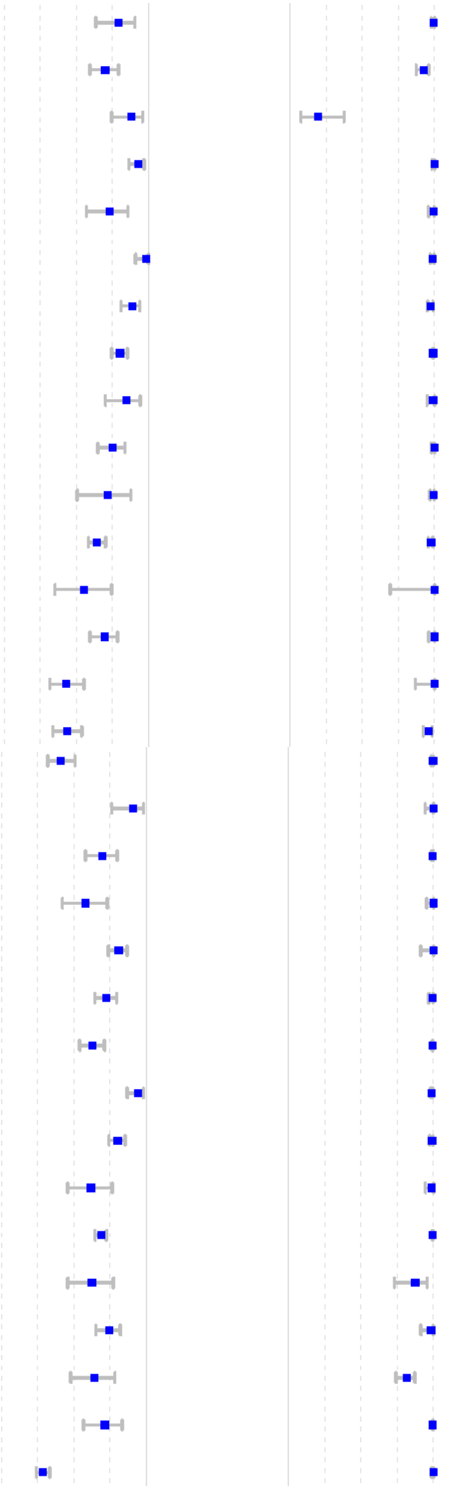

| | | | | | | | |
|---|---|---|---|---|---|---|---|
| Dankova, f142.1 | 1227 | 14 | 23 | 1190 | 0 | 0.38 [0.22, 0.55] | 1.00 [1.00, 1.00] |
| Van Honacker, f143.5 | 98 | 48 | 10 | 40 | 0 | 0.83 [0.71, 0.91] | 1.00 [0.91, 1.00] |
| Van Honacker, f143.6 | 4195 | 200 | 169 | 3814 | 12 | 0.54 [0.49, 0.59] | 1.00 [1.00, 1.00] |
| Wertenauer, f150.1 | 2215 | 204 | 134 | 1872 | 5 | 0.60 [0.55, 0.66] | 1.00 [0.99, 1.00] |
| Kim, f155.1 | 165 | 58 | 7 | 96 | 4 | 0.89 [0.79, 0.96] | 0.96 [0.90, 0.99] |
| Osterman, f157.1 | 409 | 25 | 81 | 303 | 0 | 0.24 [0.16, 0.33] | 1.00 [0.99, 1.00] |
| Jegerlehner, f172.1 | 1462 | 92 | 49 | 1319 | 2 | 0.65 [0.57, 0.73] | 1.00 [1.00, 1.00] |
| FINDdx, f181.1 | 333 | 59 | 50 | 218 | 6 | 0.54 [0.44, 0.64] | 0.97 [0.94, 0.99] |
| FINDdx, f181.2 | 335 | 71 | 29 | 234 | 1 | 0.71 [0.61, 0.80] | 1.00 [0.98, 1.00] |
| FINDdx, f182.1 | 214 | 66 | 12 | 135 | 1 | 0.85 [0.75, 0.92] | 0.99 [0.96, 1.00] |
| Korenkov, f187.1 | 1849 | 89 | 101 | 1657 | 2 | 0.47 [0.40, 0.54] | 1.00 [1.00, 1.00] |
| Fourati, f190.2 | 634 | 180 | 117 | 314 | 23 | 0.61 [0.55, 0.66] | 0.93 [0.90, 0.96] |

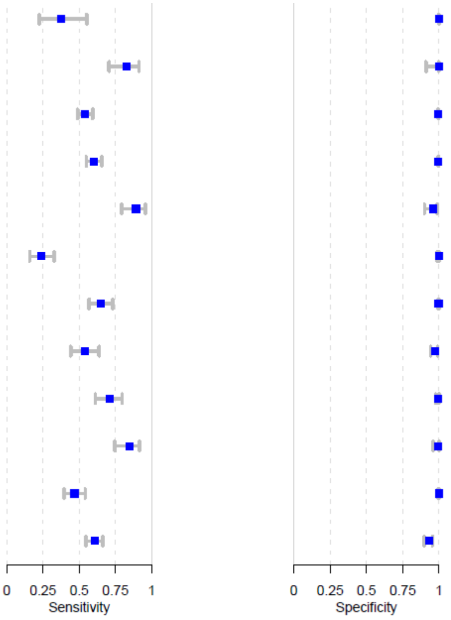


**Standard F**

| Author, Study ID | Sample size | TP | FN | TN | FP | Sensitivity [95%CI] | Specificity [95%CI] |
|---|---|---|---|---|---|---|---|
| Drevinek, a10.2 | 591 | 139 | 84 | 366 | 2 | 0.62 [0.56, 0.69] | 1.00 [0.98, 1.00] |
| Liotti, a22.1 | 359 | 49 | 55 | 251 | 4 | 0.47 [0.37, 0.57] | 0.98 [0.96, 1.00] |
| Porte, a32.2 | 64 | 29 | 3 | 31 | 1 | 0.91 [0.75, 0.98] | 0.97 [0.84, 1.00] |
| FINDdx, a63.1 | 453 | 93 | 27 | 326 | 7 | 0.78 [0.69, 0.85] | 0.98 [0.96, 0.99] |
| FINDdx, a63.2 | 676 | 27 | 12 | 617 | 20 | 0.69 [0.52, 0.83] | 0.97 [0.95, 0.98] |
| Osterman, f20.1 | 549 | 115 | 74 | 352 | 8 | 0.61 [0.54, 0.68] | 0.98 [0.96, 0.99] |
| Pérez...García, f111.2 | 356 | 113 | 57 | 181 | 5 | 0.66 [0.59, 0.74] | 0.97 [0.94, 0.99] |
| Menchinelli, f145.1 | 2898 | 137 | 63 | 2617 | 81 | 0.69 [0.62, 0.75] | 0.97 [0.96, 0.98] |
| Baccani, f165.2 | 93 | 10 | 18 | 65 | 0 | 0.36 [0.19, 0.56] | 1.00 [0.94, 1.00] |
| Kahn, f167.1 | 3110 | 57 | 39 | 2983 | 31 | 0.59 [0.49, 0.69] | 0.99 [0.98, 0.99] |
| Orsi, f191.2 | 110 | 52 | 8 | 50 | 0 | 0.87 [0.75, 0.94] | 1.00 [0.93, 1.00] |

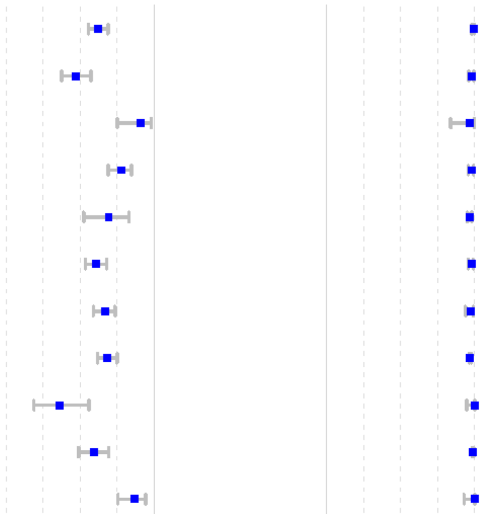

**Coris**

| Author, Study ID | Sample size | TP | FN | TN | FP | Sensitivity [95%CI] | Specificity [95%CI] |
|---|---|---|---|---|---|---|---|
| Lambert−Niclot, a18.1 | 138 | 47 | 47 | 44 | 0 | 0.50 [0.40, 0.60] | 1.00 [0.92, 1.00] |
| Scohy, a34.1 | 148 | 32 | 74 | 42 | 0 | 0.30 [0.22, 0.40] | 1.00 [0.92, 1.00] |
| Mertens, a48.1 | 328 | 76 | 56 | 195 | 1 | 0.58 [0.49, 0.66] | 1.00 [0.97, 1.00] |
| Ciotti, f24.1 | 50 | 12 | 27 | 11 | 0 | 0.31 [0.17, 0.48] | 1.00 [0.72, 1.00] |
| Seynaeve, f137.2 | 100 | 31 | 19 | 50 | 0 | 0.62 [0.47, 0.75] | 1.00 [0.93, 1.00] |
| Kanaujia, f162.1 | 484 | 136 | 53 | 293 | 2 | 0.72 [0.65, 0.78] | 0.99 [0.98, 1.00] |
| Fourati, f190.1 | 634 | 105 | 192 | 337 | 0 | 0.35 [0.30, 0.41] | 1.00 [0.99, 1.00] |

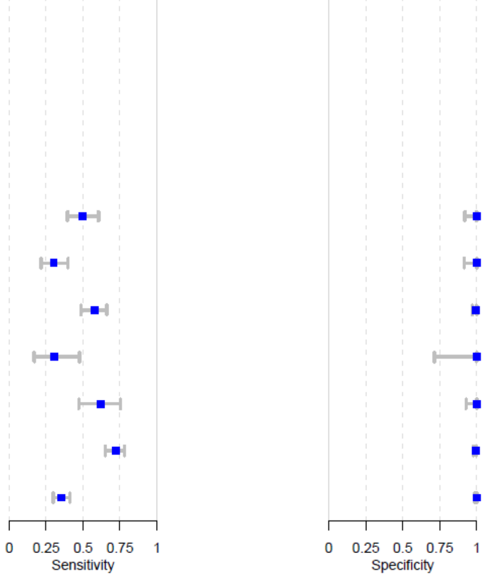

**Stark**

| Author, Study ID | Sample size | TP | FN | TN | FP | Sensitivity [95%CI] | Specificity [95%CI] |
|---|---|---|---|---|---|---|---|
| Di Domenico, f140.1 | 433 | 36 | 0 | 382 | 15 | 1.00 [0.90, 1.00] | 0.96 [0.94, 0.98] |

**Biosynex**

| Author, Study ID | Sample size | TP | FN | TN | FP | Sensitivity [95%CI] | Specificity [95%CI] |
|---|---|---|---|---|---|---|---|
| Van Honacker, f143.1 | 97 | 52 | 6 | 18 | 21 | 0.90 [0.79, 0.96] | 0.46 [0.30, 0.63] |
| Jung, f163.1 | 308 | 29 | 4 | 271 | 4 | 0.88 [0.72, 0.97] | 0.98 [0.96, 1.00] |
| Fourati, f190.6 | 634 | 178 | 119 | 337 | 0 | 0.60 [0.54, 0.66] | 1.00 [0.99, 1.00] |

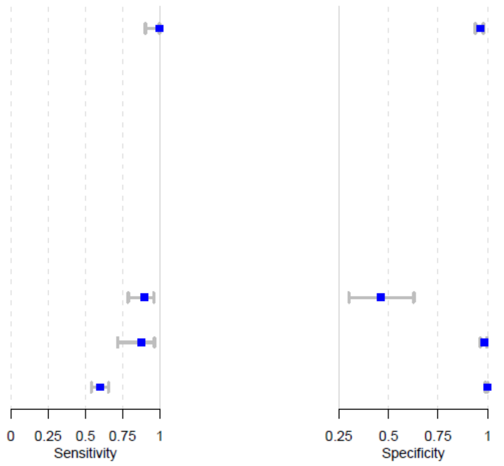


**SureScreen F**

| Author, Study ID | Sample size | TP | FN | TN | FP | Sensitivity [95%CI] | Specificity [95%CI] |
|---|---|---|---|---|---|---|---|
| Pickering, f73.6 | 200 | 69 | 31 | 98 | 2 | 0.69 [0.59, 0.78] | 0.98 [0.93, 1.00] |

**Dräger**

| Author, Study ID | Sample size | TP | FN | TN | FP | Sensitivity [95%CI] | Specificity [95%CI] |
|---|---|---|---|---|---|---|---|
| Osmanodja, f79.1 | 379 | 62 | 8 | 308 | 1 | 0.89 [0.79, 0.95] | 1.00 [0.98, 1.00] |

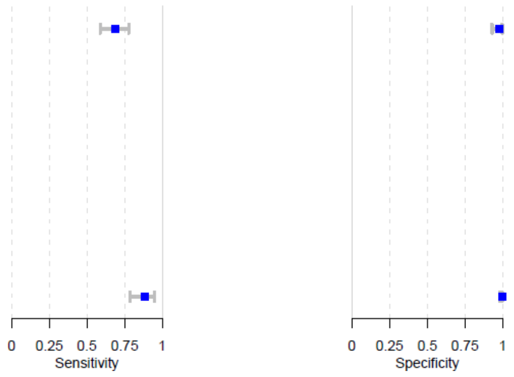


**SureScreen V**

| Author, Study ID | Sample size | TP | FN | TN | FP | Sensitivity [95%CI] | Specificity [95%CI] |
|---|---|---|---|---|---|---|---|
| Baro, f33.5 | 286 | 29 | 72 | 181 | 4 | 0.29 [0.20, 0.39] | 0.98 [0.95, 0.99] |
| Pickering, f73.3 | 200 | 65 | 35 | 100 | 0 | 0.65 [0.55, 0.74] | 1.00 [0.96, 1.00] |

**Lepu**

| Author, Study ID | Sample size | TP | FN | TN | FP | Sensitivity [95%CI] | Specificity [95%CI] |
|---|---|---|---|---|---|---|---|
| Baro, f33.4 | 286 | 46 | 55 | 165 | 20 | 0.46 [0.36, 0.56] | 0.89 [0.84, 0.93] |

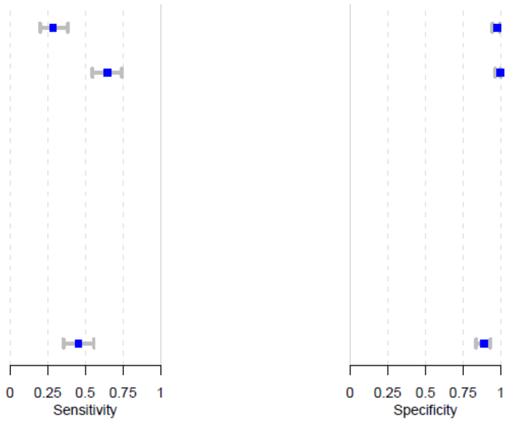

**V−Chek**

| Author, Study ID | Sample size | TP | FN | TN | FP | Sensitivity [95%CI] | Specificity [95%CI] |
|---|---|---|---|---|---|---|---|
| Kipritci, f146.1 | 110 | 21 | 13 | 76 | 0 | 0.62 [0.44, 0.78] | 1.00 [0.95, 1.00] |

**Flowflex FIA**

| Author, Study ID | Sample size | TP | FN | TN | FP | Sensitivity [95%CI] | Specificity [95%CI] |
|---|---|---|---|---|---|---|---|
| Karon, f148.3 | 347 | 174 | 23 | 150 | 0 | 0.88 [0.83, 0.92] | 1.00 [0.98, 1.00] |

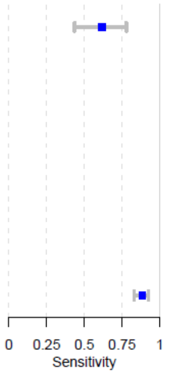


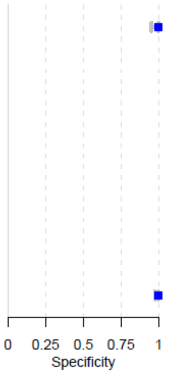


**VIRO AAZ**

| Author, Study ID | Sample size | TP | FN | TN | FP | Sensitivity [95%CI] | Specificity [95%CI] |
|---|---|---|---|---|---|---|---|
| Schwob, a35.3 | 324 | 116 | 22 | 186 | 0 | 0.84 [0.77, 0.90] | 1.00 [0.98, 1.00] |
| Cassuto, f149.1 | 234 | 31 | 1 | 202 | 0 | 0.97 [0.84, 1.00] | 1.00 [0.98, 1.00] |
| Fourati, f190.4 | 634 | 183 | 114 | 337 | 0 | 0.62 [0.56, 0.67] | 1.00 [0.99, 1.00] |

**Savant**

| Author, Study ID | Sample size | TP | FN | TN | FP | Sensitivity [95%CI] | Specificity [95%CI] |
|---|---|---|---|---|---|---|---|
| Weitzel, a41.3 | 109 | 13 | 65 | 31 | 0 | 0.17 [0.09, 0.27] | 1.00 [0.89, 1.00] |

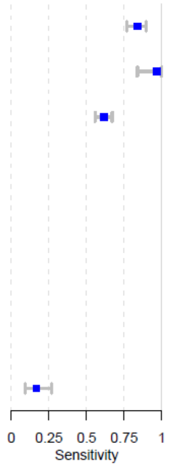


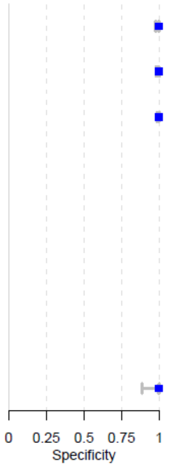


**VIVADiag**

| Author, Study ID | Sample size | TP | FN | TN | FP | Sensitivity [95%CI] | Specificity [95%CI] |
|---|---|---|---|---|---|---|---|
| Homza, f87.5 | 268 | 38 | 53 | 170 | 7 | 0.42 [0.32, 0.53] | 0.96 [0.92, 0.98] |

**ACCUCARE**

| Author, Study ID | Sample size | TP | FN | TN | FP | Sensitivity [95%CI] | Specificity [95%CI] |
|---|---|---|---|---|---|---|---|
| Thakur, f88.1 | 677 | 29 | 55 | 592 | 1 | 0.34 [0.24, 0.46] | 1.00 [0.99, 1.00] |

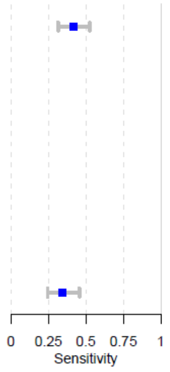


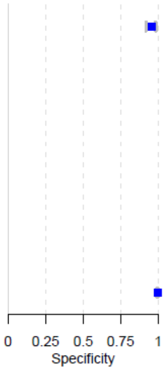
