## Supplementary figures and images for "Accuracy of rapid point-of-care antigen-based diagnostics for SARS-CoV-2: an updated systematic review and meta-analysis with meta regression analyzing influencing factors"

### S2 Fig

#### S2 Fig. Details of QUADAS assessment.

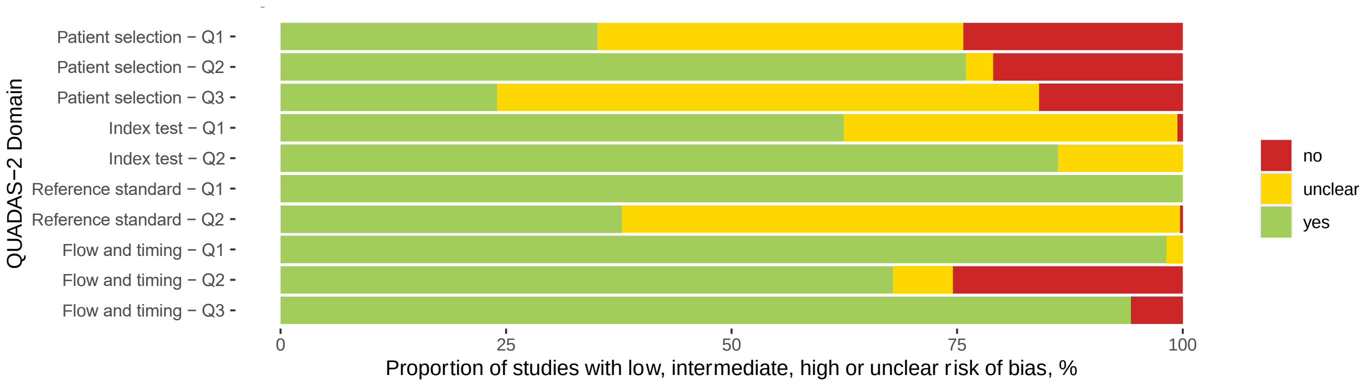
