## Supplementary material for "Accuracy of rapid point-of-care antigen-based diagnostics for SARS-CoV-2: an updated systematic review and meta-analysis with meta regression analyzing influencing factors": S2 Text

S2 Text. Search strategy.

### Review questions

To assess the accuracy and ease-of-use of marketable antigen point of care diagnostics for SARS-CoV-2 compared to RT-PCR based on manufacturer independent evaluations.

Restriction: start Dezember 2019

### Definitions

P

SARS-CoV-2

I

Antigen nachweisenden Schnelltests Ag RDT

### Strategie

| 1 | P |
| --- | --- |
| 2 | I |
| 3 | 1 AND 2 |

### Searched Databases

- PubMed
- Web of Science Core Collection
- BioRxiv
- MedRxiv

### PubMed

P

(**"Severe Acute Respiratory Syndrome Coronavirus 2"[Supplementary Concept] OR**

**"COVID-19" [Supplementary Concept] OR**

**"Betacoronavirus"[Mesh] OR**

**"Coronavirus"[Mesh] OR**

covid*[tw] OR

"coronavirus*"[tw] OR

"corona virus*"[tw] OR

ncov*[tw] OR

"n cov*"[tw] OR

sarscov*[tw] OR

"sars cov*"[tw] OR

"2019nCoV*"[tw] OR

"2019 nCoV*"[tw] OR

"sars2*"[tw] OR

"sars 2*"[tw])

I

**"Point-of-Care Testing"[Mesh] OR**

Antigen[tw] OR

“Lateral flow”[tw] OR

RDT[tw] OR

(("Point of Care*"[tw] OR

"Bedside*"[tw] OR

Rapid*[tw])

AND

Test*[tw])

**P**

("Severe Acute Respiratory Syndrome Coronavirus 2"[Supplementary Concept] OR "COVID-19"[Supplementary Concept] OR "Betacoronavirus"[MeSH Terms] OR "Coronavirus"[MeSH Terms] OR "covid*"[Text Word] OR "coronavirus*"[Text Word] OR "corona virus*"[Text Word] OR "ncov*"[Text Word] OR "n cov*"[Text Word] OR "sarscov*"[Text Word] OR "sars cov*"[Text Word] OR "2019ncov*"[Text Word] OR "2019 ncov*"[Text Word] OR "sars2*"[Text Word] OR "sars 2*"[Text Word])

**I**

("Point-of-Care Testing"[MeSH Terms] OR "antigen"[Text Word] OR "lateral flow"[Text Word] OR "RDT"[Text Word] OR (("point of care*"[Text Word] OR "bedside*"[Text Word] OR "rapid*"[Text Word]) AND "test*"[Text Word]))

2019/12/01:2021/08/31[Date - Publication]

### Web of Science Core Collection

P

"covid*" OR

"coronavirus*" OR

"corona virus*" OR

"ncov*" OR

"n cov*" OR

"sarscov*" OR

"sars cov*" OR

"2019nCoV*" OR

"2019 nCoV*" OR

"sars2*" OR

"sars 2*"

I

"antigen" OR

"Lateral flow" OR

"RDT" OR

(("Point of Care*" OR

"Bedside*" OR

"Rapid*")

AND

Test*))

P

TS=("covid*" OR "coronavirus*" OR "corona virus*" OR "ncov*" OR "n cov*" OR "sarscov*" OR "sars cov*" OR "2019nCoV*" OR "2019 nCoV*" OR "sars2*" OR "sars 2*")

I

TS=("antigen" OR "Lateral flow" OR "RDT" OR (("Point of Care*" OR "Bedside*" OR "Rapid*")AND Test*))

Filter (year to date 2021/08/31)

### Bio_MedRxiv

https://europepmc.org/

P

Covid* OR

Coronavirus* OR

"corona virus*" OR

Ncov* OR

"n cov*" OR

Sarscov* OR

"Sars cov*" OR

2019nCoV* OR

"2019 nCoV*" OR

sars2* OR

"sars 2*"

I

"Antigen*" OR

"Lateral flow*" OR

"RDT" OR

"Rapid test*" OR

"Bedside*" OR

"Point of Care*"

**P AND I**

(covid* OR Coronavirus* OR "corona virus*" OR ncov* OR "n cov*" OR sarscov* OR "sars cov*" OR 2019nCov* OR "2019 nCov*" OR sars2* OR "sars 2*")

AND

(Antigen OR "Lateral flow" OR RDT OR "Rapid test*" OR Bedside* OR "Point of Care*")

AND

(PUBLISHER:MedRxiv OR PUBLISHER:BioRxiv)

AND

FIRST_PDATE:[2019-12-01 TO 2021-08-31]
