## Supplementary material for "Accuracy of rapid point-of-care antigen-based diagnostics for SARS-CoV-2: an updated systematic review and meta-analysis with meta regression analyzing influencing factors": S3 Fig

<u>S3 Fig. Forest plots for subgroup analysis by Ct-values.</u>

Caption: CI = confidence interval

#### Fig A – Forest plots for Ct-values lower 20

| Author, Study ID | Sample size | Sensitivity [95%CI] |
|---|---|---|
| Alemany, a02.ov | 258 | 0.98 [0.96, 1.00] |
| Gupta, a13.1 | 5 | 0.80 [0.28, 1.00] |
| Gupta, a13.1 | 27 | 0.96 [0.81, 1.00] |
| Krüttgen, a16.1 | 5 | 1.00 [0.48, 1.00] |
| Takeda, a50.1 | 9 | 1.00 [0.66, 1.00] |
| Favresse, f31.1 | 2 | 1.00 [0.16, 1.00] |
| Favresse, f31.1 | 34 | 0.97 [0.85, 1.00] |
| Favresse, f31.2 | 2 | 0.50 [0.01, 0.99] |
| Favresse, f31.2 | 34 | 0.94 [0.80, 0.99] |
| Favresse, f31.3 | 2 | 1.00 [0.16, 1.00] |
| Favresse, f31.3 | 34 | 0.97 [0.85, 1.00] |
| Favresse, f31.4 | 2 | 1.00 [0.16, 1.00] |
| Favresse, f31.4 | 34 | 0.97 [0.85, 1.00] |
| Pena, f36.1 | 31 | 0.97 [0.83, 1.00] |
| Young, f56.1 | 71 | 0.83 [0.72, 0.91] |
| Young, f56.1 | 62 | 0.76 [0.63, 0.86] |
| Faíco−Filho, f63.1 | 13 | 1.00 [0.75, 1.00] |
| Bouassa, f67.1 | 25 | 1.00 [0.86, 1.00] |
| Thell, f81.1 | 51 | 1.00 [0.93, 1.00] |
| Sberna, f83.1 | 4 | 0.75 [0.19, 0.99] |
| Pérez...García, f111.1 | 41 | 1.00 [0.91, 1.00] |
| Pérez...García, f111.2 | 41 | 1.00 [0.91, 1.00] |
| Blairon, f113.1 | 28 | 1.00 [0.88, 1.00] |
| Blairon, f113.2 | 28 | 1.00 [0.88, 1.00] |
| Blairon, f113.3 | 28 | 1.00 [0.88, 1.00] |
| Nordgren, f117.1 | 47 | 0.94 [0.82, 0.99] |
| Nordgren, f117.2 | 47 | 0.98 [0.89, 1.00] |
| Fernandez−Montero, f126.1 | 9 | 1.00 [0.66, 1.00] |
| García−Fiñana, f131.1 | 11 | 0.91 [0.59, 1.00] |
| Lee, f136.1 | 44 | 0.89 [0.75, 0.96] |
| Carbonell−Sahuquillo, f141.1 | 17 | 1.00 [0.80, 1.00] |
| Van Honacker, f143.1 | 14 | 1.00 [0.77, 1.00] |
| Van Honacker, f143.2 | 14 | 1.00 [0.77, 1.00] |
| Van Honacker, f143.3 | 14 | 1.00 [0.77, 1.00] |
| Van Honacker, f143.4 | 13 | 1.00 [0.75, 1.00] |
| Van Honacker, f143.5 | 14 | 1.00 [0.77, 1.00] |
| Van Honacker, f143.6 | 61 | 1.00 [0.94, 1.00] |
| Kurihara, f152.1 | 16 | 1.00 [0.79, 1.00] |
| Kolwijck, f168.1 | 25 | 1.00 [0.86, 1.00] |
| Mayanskiy, f188.1 | 26 | 1.00 [0.87, 1.00] |
| Fourati, f190.1 | 40 | 0.90 [0.76, 0.97] |
| Fourati, f190.2 | 40 | 1.00 [0.91, 1.00] |
| Fourati, f190.3 | 40 | 0.95 [0.83, 0.99] |
| Fourati, f190.4 | 40 | 1.00 [0.91, 1.00] |
| Fourati, f190.5 | 40 | 0.80 [0.64, 0.91] |
| Fourati, f190.6 | 40 | 0.98 [0.87, 1.00] |
| Suzuki, f194.1 | 28 | 0.96 [0.82, 1.00] |
| Suzuki, f194.2 | 28 | 1.00 [0.88, 1.00] |
| Thirion−Romero, f195.1 | 75 | 0.88 [0.78, 0.94] |

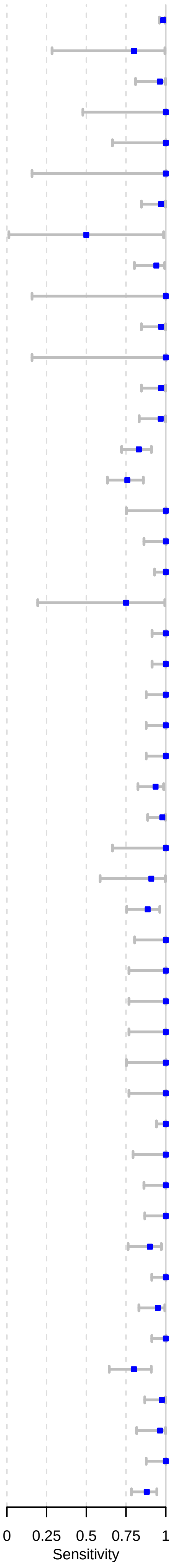

Fig B – Forest plots for Ct-values greater 20

| Author, Study ID | Sample size | Sensitivity [95%CI] |
|---|---|---|
| Alemany, a02.ov | 305 | 0.99 [0.98, 1.00] |
| Gupta, a13.1 | 28 | 0.86 [0.67, 0.96] |
| Krüttgen, a16.1 | 12 | 1.00 [0.74, 1.00] |
| Takeda, a50.1 | 23 | 1.00 [0.85, 1.00] |
| Favresse, f31.1 | 22 | 0.86 [0.65, 0.97] |
| Favresse, f31.2 | 22 | 0.96 [0.77, 1.00] |
| Favresse, f31.3 | 22 | 0.96 [0.77, 1.00] |
| Favresse, f31.4 | 22 | 0.96 [0.77, 1.00] |
| Young, f56.1 | 54 | 0.43 [0.29, 0.57] |
| Bouassa, f67.1 | 52 | 0.94 [0.84, 0.99] |
| Sberna, f83.1 | 14 | 0.07 [0.00, 0.34] |
| Pérez...García, f111.1 | 43 | 0.93 [0.81, 0.98] |
| Pérez...García, f111.2 | 43 | 0.95 [0.84, 0.99] |
| Nordgren, f117.1 | 66 | 0.85 [0.74, 0.92] |
| Nordgren, f117.2 | 66 | 0.91 [0.81, 0.97] |
| Fernandez−Montero, f126.1 | 23 | 0.91 [0.72, 0.99] |
| Lee, f136.1 | 65 | 0.55 [0.42, 0.68] |
| Van Honacker, f143.1 | 18 | 1.00 [0.81, 1.00] |
| Van Honacker, f143.2 | 18 | 0.89 [0.65, 0.99] |
| Van Honacker, f143.3 | 18 | 1.00 [0.81, 1.00] |
| Van Honacker, f143.4 | 18 | 0.94 [0.73, 1.00] |
| Van Honacker, f143.5 | 18 | 1.00 [0.81, 1.00] |
| Van Honacker, f143.6 | 108 | 0.93 [0.86, 0.97] |
| Kurihara, f152.1 | 29 | 0.97 [0.82, 1.00] |
| Mayanskiy, f188.1 | 62 | 0.98 [0.91, 1.00] |
| Fourati, f190.1 | 90 | 0.63 [0.52, 0.73] |
| Fourati, f190.2 | 90 | 0.90 [0.82, 0.95] |
| Fourati, f190.3 | 90 | 0.83 [0.74, 0.90] |
| Fourati, f190.4 | 90 | 0.94 [0.88, 0.98] |
| Fourati, f190.5 | 90 | 0.54 [0.44, 0.65] |
| Fourati, f190.6 | 90 | 0.92 [0.85, 0.97] |
| Suzuki, f194.1 | 32 | 0.81 [0.64, 0.93] |
| Suzuki, f194.2 | 32 | 0.84 [0.67, 0.95] |
| Thirion−Romero, f195.1 | 92 | 0.73 [0.63, 0.82] |

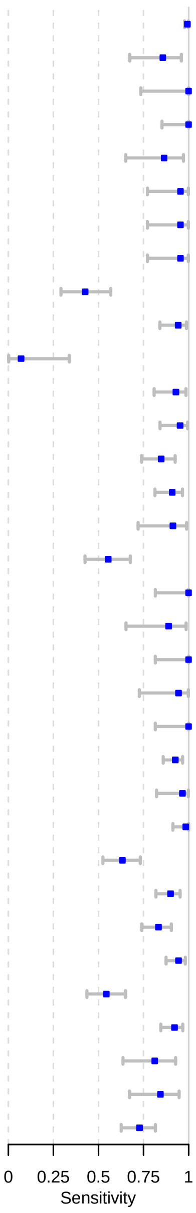

### Fig C – Forest plots for Ct-values lower 25

| Author, Study ID | Sample size | Sensitivity [95%CI] |
|---|---|---|
| Alemany, a02.ov | 305 | 0.99 [0.98, 1.00] |
| Bulilete, a07.1 | 95 | 0.90 [0.81, 0.95] |
| Gupta, a13.1 | 28 | 0.86 [0.67, 0.96] |
| Krüttgen, a16.1 | 12 | 1.00 [0.74, 1.00] |
| Krüger, a17.1 | 18 | 1.00 [0.81, 1.00] |
| Porte, a31.1 | 52 | 1.00 [0.93, 1.00] |
| Porte, a32.1 | 27 | 1.00 [0.87, 1.00] |
| Porte, a32.2 | 27 | 1.00 [0.87, 1.00] |
| Scohy, a34.1 | 10 | 1.00 [0.69, 1.00] |
| Weitzel, a41.1 | 53 | 0.85 [0.72, 0.93] |
| Weitzel, a41.3 | 52 | 0.21 [0.11, 0.35] |
| Weitzel, a41.4 | 54 | 1.00 [0.93, 1.00] |
| Takeda, a50.1 | 23 | 1.00 [0.85, 1.00] |
| Krüger, a52.ov | 72 | 0.96 [0.88, 0.99] |
| Agullo, a56.2 | 68 | 0.75 [0.63, 0.85] |
| Agullo, a56.3 | 63 | 0.38 [0.26, 0.51] |
| Strömer, f11.1 | 32 | 1.00 [0.89, 1.00] |
| Pollock, f17.1 | 150 | 0.99 [0.96, 1.00] |
| Favresse, f31.1 | 22 | 0.86 [0.65, 0.97] |
| Favresse, f31.2 | 22 | 0.96 [0.77, 1.00] |
| Favresse, f31.3 | 22 | 0.96 [0.77, 1.00] |
| Favresse, f31.4 | 22 | 0.96 [0.77, 1.00] |
| Pena, f36.1 | 30 | 0.67 [0.47, 0.83] |
| Jääskeläinen, f50.1 | 89 | 0.99 [0.94, 1.00] |
| Jääskeläinen, f50.2 | 97 | 0.99 [0.94, 1.00] |
| Jääskeläinen, f50.3 | 92 | 0.98 [0.92, 1.00] |
| Pérez−García, f52.1 | 84 | 0.94 [0.87, 0.98] |
| Pérez−García, f52.2 | 84 | 0.96 [0.90, 0.99] |
| Young, f56.1 | 54 | 0.43 [0.29, 0.57] |
| Krüger, f58.1 | 95 | 0.93 [0.85, 0.97] |
| Pollock, f59.1 | 95 | 0.96 [0.90, 0.99] |
| Pollock, f59.1 | 12 | 1.00 [0.74, 1.00] |
| Faíco−Filho, f63.1 | 37 | 0.97 [0.86, 1.00] |
| Pickering, f73.1 | 69 | 0.99 [0.92, 1.00] |
| Pickering, f73.3 | 69 | 0.91 [0.82, 0.97] |
| Pickering, f73.4 | 69 | 0.96 [0.88, 0.99] |
| Pickering, f73.5 | 69 | 0.94 [0.86, 0.98] |
| Pickering, f73.6 | 69 | 0.88 [0.78, 0.95] |
| Pickering, f73.7 | 82 | 0.84 [0.74, 0.91] |
| Pickering, f73.8 | 66 | 0.97 [0.90, 1.00] |
| Pickering, f73.9 | 52 | 0.96 [0.87, 1.00] |
| Thell, f81.1 | 125 | 0.98 [0.94, 1.00] |
| Sberna, f83.1 | 14 | 0.07 [0.00, 0.34] |
| Thakur, f88.1 | 60 | 0.45 [0.32, 0.58] |
| Pérez...García, f111.1 | 43 | 0.93 [0.81, 0.98] |
| Pérez...García, f111.2 | 43 | 0.95 [0.84, 0.99] |
| Bornemann, f112.1 | 43 | 0.95 [0.84, 0.99] |
| Blairon, f113.1 | 69 | 0.96 [0.88, 0.99] |
| Blairon, f113.2 | 69 | 0.97 [0.90, 1.00] |
| Blairon, f113.3 | 69 | 0.97 [0.90, 1.00] |
| Nordgren, f117.1 | 66 | 0.85 [0.74, 0.92] |
| Nordgren, f117.2 | 66 | 0.91 [0.81, 0.97] |
| Leixner, f125.1 | 33 | 1.00 [0.89, 1.00] |
| Fernandez−Montero, f126.1 | 23 | 0.91 [0.72, 0.99] |
| Terpos, f130.1 | 70 | 1.00 [0.95, 1.00] |
| García−Fiñana, f131.1 | 25 | 0.60 [0.39, 0.79] |
| Lee, f136.1 | 65 | 0.55 [0.42, 0.68] |
| Carbonell−Sahuquillo, f141.1 | 25 | 0.96 [0.80, 1.00] |
| Van Honacker, f143.1 | 18 | 1.00 [0.81, 1.00] |
| Van Honacker, f143.2 | 18 | 0.89 [0.65, 0.99] |
| Van Honacker, f143.3 | 18 | 1.00 [0.81, 1.00] |
| Van Honacker, f143.4 | 18 | 0.94 [0.73, 1.00] |
| Van Honacker, f143.5 | 18 | 1.00 [0.81, 1.00] |
| Van Honacker, f143.6 | 108 | 0.93 [0.86, 0.97] |
| Kurihara, f152.1 | 29 | 0.97 [0.82, 1.00] |
| Baccani, f165.2 | 5 | 1.00 [0.48, 1.00] |
| Baccani, f165.3 | 5 | 1.00 [0.48, 1.00] |
| Kolwijck, f168.1 | 34 | 1.00 [0.90, 1.00] |
| FINDdx, f175.1 | 48 | 1.00 [0.93, 1.00] |
| FINDdx, f176.1 | 29 | 1.00 [0.88, 1.00] |
| FINDdx, f177.1 | 64 | 0.70 [0.58, 0.81] |
| FINDdx, f177.2 | 33 | 0.88 [0.72, 0.97] |
| FINDdx, f178.1 | 89 | 0.96 [0.89, 0.99] |
| FINDdx, f179.1 | 37 | 0.92 [0.78, 0.98] |
| FINDdx, f180.1 | 80 | 0.98 [0.91, 1.00] |
| FINDdx, f180.2 | 81 | 0.88 [0.78, 0.94] |
| FINDdx, f181.1 | 47 | 0.89 [0.77, 0.96] |
| FINDdx, f181.2 | 52 | 0.96 [0.87, 1.00] |
| FINDdx, f182.1 | 46 | 1.00 [0.92, 1.00] |
| FINDdx, f182.2 | 46 | 1.00 [0.92, 1.00] |
| Kim, f186.1 | 15 | 1.00 [0.78, 1.00] |
| Kim, f186.2 | 54 | 0.91 [0.80, 0.97] |
| Mayanskiy, f188.1 | 62 | 0.98 [0.91, 1.00] |
| Fourati, f190.1 | 90 | 0.63 [0.52, 0.73] |
| Fourati, f190.1 | 130 | 0.71 [0.62, 0.78] |
| Fourati, f190.2 | 90 | 0.90 [0.82, 0.95] |
| Fourati, f190.2 | 130 | 0.93 [0.87, 0.97] |
| Fourati, f190.3 | 90 | 0.83 [0.74, 0.90] |
| Fourati, f190.3 | 130 | 0.87 [0.80, 0.92] |
| Fourati, f190.4 | 90 | 0.94 [0.88, 0.98] |
| Fourati, f190.4 | 130 | 0.96 [0.91, 0.99] |
| Fourati, f190.5 | 90 | 0.54 [0.44, 0.65] |
| Fourati, f190.5 | 130 | 0.62 [0.53, 0.71] |
| Fourati, f190.6 | 90 | 0.92 [0.85, 0.97] |
| Fourati, f190.6 | 130 | 0.94 [0.88, 0.97] |
| Orsi, f191.1 | 23 | 1.00 [0.85, 1.00] |
| Orsi, f191.2 | 23 | 1.00 [0.85, 1.00] |
| Thirion−Romero, f195.1 | 194 | 0.82 [0.76, 0.87] |
| Thirion−Romero, f195.1 | 92 | 0.73 [0.63, 0.82] |

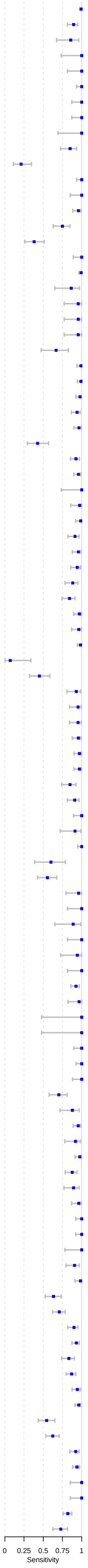

### Fig D – Forest plots for Ct-values greater 25

| Author, Study ID | Sample size | Sensitivity [95%CI] |
|---|---|---|
| Alemany, a02.ov | 285 | 0.90 [0.86, 0.93] |
| Bulilete, a07.1 | 25 | 0.40 [0.21, 0.61] |
| Gupta, a13.1 | 13 | 0.62 [0.32, 0.86] |
| Krüger, a17.1 | 29 | 0.62 [0.42, 0.79] |
| Porte, a31.1 | 18 | 0.72 [0.47, 0.90] |
| Weitzel, a41.1 | 26 | 0.15 [0.04, 0.35] |
| Weitzel, a41.3 | 26 | 0.08 [0.01, 0.25] |
| Weitzel, a41.4 | 26 | 0.54 [0.33, 0.73] |
| Takeda, a50.1 | 18 | 0.89 [0.65, 0.99] |
| Krüger, a52.ov | 33 | 0.67 [0.48, 0.82] |
| Favresse, f31.1 | 19 | 0.32 [0.13, 0.57] |
| Favresse, f31.2 | 19 | 0.53 [0.29, 0.76] |
| Favresse, f31.3 | 19 | 0.68 [0.43, 0.87] |
| Favresse, f31.4 | 19 | 0.53 [0.29, 0.76] |
| Jääskeläinen, f50.1 | 34 | 0.82 [0.66, 0.93] |
| Jääskeläinen, f50.2 | 35 | 0.69 [0.51, 0.83] |
| Jääskeläinen, f50.3 | 34 | 0.76 [0.59, 0.89] |
| Pérez−García, f52.1 | 86 | 0.14 [0.07, 0.23] |
| Pérez−García, f52.2 | 86 | 0.24 [0.16, 0.35] |
| Young, f56.1 | 23 | 0.17 [0.05, 0.39] |
| Krüger, f58.1 | 51 | 0.63 [0.48, 0.76] |
| Sberna, f83.1 | 23 | 0.04 [0.00, 0.22] |
| Thakur, f88.1 | 21 | 0.10 [0.01, 0.30] |
| Pérez...García, f111.1 | 46 | 0.41 [0.27, 0.57] |
| Pérez...García, f111.2 | 46 | 0.52 [0.37, 0.67] |
| Nordgren, f117.1 | 20 | 0.45 [0.23, 0.69] |
| Nordgren, f117.2 | 20 | 0.55 [0.32, 0.77] |
| Fernandez−Montero, f126.1 | 9 | 0.44 [0.14, 0.79] |
| García−Fiñana, f131.1 | 17 | 0.18 [0.04, 0.43] |
| Lee, f136.1 | 118 | 0.17 [0.11, 0.25] |
| Van Honacker, f143.1 | 18 | 0.89 [0.65, 0.99] |
| Van Honacker, f143.2 | 18 | 0.44 [0.22, 0.69] |
| Van Honacker, f143.3 | 18 | 0.72 [0.47, 0.90] |
| Van Honacker, f143.4 | 18 | 0.72 [0.47, 0.90] |
| Van Honacker, f143.5 | 18 | 0.72 [0.47, 0.90] |
| Van Honacker, f143.6 | 64 | 0.55 [0.42, 0.67] |
| Kurihara, f152.1 | 13 | 1.00 [0.75, 1.00] |
| Baccani, f165.2 | 9 | 0.56 [0.21, 0.86] |
| Baccani, f165.3 | 9 | 0.44 [0.14, 0.79] |
| Kim, f186.1 | 12 | 0.92 [0.62, 1.00] |
| Kim, f186.2 | 45 | 0.98 [0.88, 1.00] |
| Mayanskiy, f188.1 | 42 | 0.93 [0.80, 0.98] |
| Fourati, f190.1 | 74 | 0.15 [0.08, 0.25] |
| Fourati, f190.2 | 74 | 0.65 [0.53, 0.76] |
| Fourati, f190.3 | 74 | 0.58 [0.46, 0.70] |
| Fourati, f190.4 | 74 | 0.66 [0.54, 0.77] |
| Fourati, f190.5 | 74 | 0.19 [0.11, 0.30] |
| Fourati, f190.6 | 74 | 0.64 [0.52, 0.74] |
| Orsi, f191.1 | 32 | 0.97 [0.84, 1.00] |
| Orsi, f191.2 | 32 | 0.91 [0.75, 0.98] |
| Thirion−Romero, f195.1 | 88 | 0.33 [0.23, 0.44] |
| Thirion−Romero, f195.1 | 195 | 0.25 [0.19, 0.31] |

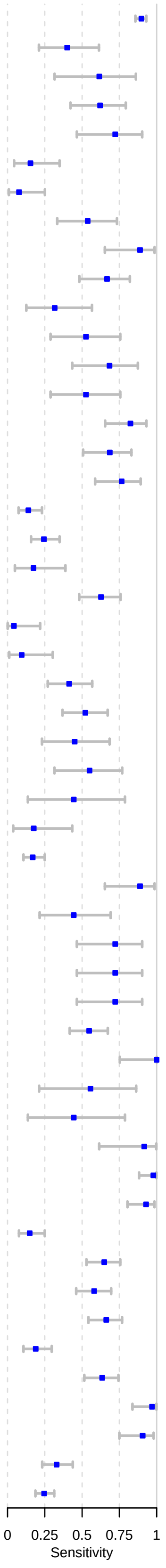

### Fig E – Forest plots for Ct-values lower 30

| Author, Study ID | Sample size | Sensitivity [95%CI] |
|---|---|---|
| Alemany, a02.ov | 285 | 0.90 [0.86, 0.93] |
| Bulilete, a07.1 | 25 | 0.40 [0.21, 0.61] |
| Gremmels, a12.1 | 106 | 0.95 [0.89, 0.98] |
| Gremmels, a12.2 | 49 | 0.98 [0.89, 1.00] |
| Gupta, a13.1 | 13 | 0.62 [0.32, 0.86] |
| Merino−Amador, a25.1 | 254 | 0.94 [0.91, 0.97] |
| Nalumansi, a27.1 | 37 | 0.92 [0.78, 0.98] |
| Pilarowski, a29.1 | 15 | 0.93 [0.68, 1.00] |
| Pilarowski, a29.1 | 16 | 0.94 [0.70, 1.00] |
| Takeda, a50.1 | 18 | 0.89 [0.65, 0.99] |
| Krüger, a52.ov | 93 | 0.94 [0.86, 0.98] |
| Agullo, a56.2 | 96 | 0.58 [0.48, 0.68] |
| Agullo, a56.3 | 87 | 0.32 [0.23, 0.43] |
| Strömer, f11.1 | 108 | 0.73 [0.64, 0.81] |
| Pollock, f17.1 | 214 | 0.96 [0.92, 0.98] |
| Favresse, f31.1 | 19 | 0.32 [0.13, 0.57] |
| Favresse, f31.2 | 19 | 0.53 [0.29, 0.76] |
| Favresse, f31.3 | 19 | 0.68 [0.43, 0.87] |
| Favresse, f31.4 | 19 | 0.53 [0.29, 0.76] |
| Jääskeläinen, f50.1 | 34 | 0.82 [0.66, 0.93] |
| Jääskeläinen, f50.1 | 123 | 0.94 [0.89, 0.98] |
| Jääskeläinen, f50.2 | 35 | 0.69 [0.51, 0.83] |
| Jääskeläinen, f50.2 | 132 | 0.91 [0.85, 0.95] |
| Jääskeläinen, f50.3 | 34 | 0.76 [0.59, 0.89] |
| Jääskeläinen, f50.3 | 126 | 0.92 [0.86, 0.96] |
| Okoye, f51.1 | 36 | 0.67 [0.49, 0.81] |
| Okoye, f51.1 | 42 | 0.57 [0.41, 0.72] |
| Young, f56.1 | 23 | 0.17 [0.05, 0.39] |
| Krüger, f58.1 | 122 | 0.90 [0.83, 0.95] |
| Pollock, f59.1 | 133 | 0.79 [0.71, 0.86] |
| Pollock, f59.1 | 29 | 0.83 [0.64, 0.94] |
| Faíco−Filho, f63.1 | 59 | 0.88 [0.77, 0.95] |
| Thell, f81.1 | 173 | 0.92 [0.88, 0.96] |
| Sberna, f83.1 | 23 | 0.04 [0.00, 0.22] |
| Thakur, f88.1 | 21 | 0.10 [0.01, 0.30] |
| Holzner, f97.1 | 456 | 0.80 [0.76, 0.84] |
| Holzner, f97.1 | 423 | 0.80 [0.76, 0.83] |
| Pérez...García, f111.1 | 46 | 0.41 [0.27, 0.57] |
| Pérez...García, f111.2 | 46 | 0.52 [0.37, 0.67] |
| Blairon, f113.1 | 97 | 0.86 [0.77, 0.92] |
| Blairon, f113.2 | 97 | 0.86 [0.77, 0.92] |
| Blairon, f113.3 | 97 | 0.87 [0.78, 0.93] |
| Nordgren, f117.1 | 20 | 0.45 [0.23, 0.69] |
| Nordgren, f117.2 | 20 | 0.55 [0.32, 0.77] |
| Leixner, f125.1 | 61 | 0.92 [0.82, 0.97] |
| Fernandez−Montero, f126.1 | 9 | 0.44 [0.14, 0.79] |
| Terpos, f130.1 | 100 | 0.99 [0.95, 1.00] |
| García−Fiñana, f131.1 | 17 | 0.18 [0.04, 0.43] |
| Lee, f136.1 | 118 | 0.17 [0.11, 0.25] |
| Seynaeve, f137.1 | 45 | 0.91 [0.79, 0.98] |
| Seynaeve, f137.2 | 45 | 0.69 [0.53, 0.82] |
| Carbonell−Sahuquillo, f141.1 | 29 | 0.83 [0.64, 0.94] |
| Van Honacker, f143.1 | 18 | 0.89 [0.65, 0.99] |
| Van Honacker, f143.2 | 18 | 0.44 [0.22, 0.69] |
| Van Honacker, f143.3 | 18 | 0.72 [0.47, 0.90] |
| Van Honacker, f143.4 | 18 | 0.72 [0.47, 0.90] |
| Van Honacker, f143.5 | 18 | 0.72 [0.47, 0.90] |
| Van Honacker, f143.6 | 64 | 0.55 [0.42, 0.67] |
| Kurihara, f152.1 | 13 | 1.00 [0.75, 1.00] |
| Kim, f155.1 | 54 | 0.94 [0.85, 0.99] |
| Tinker, f156.1 | 9 | 0.89 [0.52, 1.00] |
| Baccani, f165.2 | 9 | 0.56 [0.21, 0.86] |
| Baccani, f165.3 | 9 | 0.44 [0.14, 0.79] |
| Kahn, f167.1 | 81 | 0.64 [0.53, 0.75] |
| Kolwijck, f168.1 | 41 | 0.95 [0.84, 0.99] |
| Jegerlehner, f172.1 | 116 | 0.79 [0.71, 0.86] |
| Jegerlehner, f172.1 | 109 | 0.84 [0.76, 0.91] |
| FINDdx, f177.1 | 86 | 0.58 [0.47, 0.69] |
| FINDdx, f177.2 | 51 | 0.76 [0.62, 0.87] |
| FINDdx, f178.1 | 105 | 0.90 [0.82, 0.95] |
| FINDdx, f179.1 | 93 | 0.74 [0.64, 0.83] |
| FINDdx, f180.1 | 96 | 0.94 [0.87, 0.98] |
| FINDdx, f180.2 | 103 | 0.75 [0.65, 0.83] |
| FINDdx, f181.1 | 87 | 0.66 [0.55, 0.75] |
| FINDdx, f181.2 | 84 | 0.83 [0.74, 0.91] |
| FINDdx, f182.1 | 72 | 0.92 [0.83, 0.97] |
| FINDdx, f182.2 | 72 | 0.92 [0.83, 0.97] |
| Kim, f186.1 | 12 | 0.92 [0.62, 1.00] |
| Kim, f186.2 | 45 | 0.98 [0.88, 1.00] |
| Mayanskiy, f188.1 | 42 | 0.93 [0.80, 0.98] |
| Fourati, f190.1 | 74 | 0.15 [0.08, 0.25] |
| Fourati, f190.2 | 74 | 0.65 [0.53, 0.76] |
| Fourati, f190.3 | 74 | 0.58 [0.46, 0.70] |
| Fourati, f190.4 | 74 | 0.66 [0.54, 0.77] |
| Fourati, f190.5 | 74 | 0.19 [0.11, 0.30] |
| Fourati, f190.6 | 74 | 0.64 [0.52, 0.74] |
| Orsi, f191.1 | 32 | 0.97 [0.84, 1.00] |
| Orsi, f191.2 | 32 | 0.91 [0.75, 0.98] |
| Suzuki, f194.1 | 32 | 0.81 [0.64, 0.93] |
| Suzuki, f194.2 | 32 | 0.84 [0.67, 0.95] |
| Thirion−Romero, f195.1 | 88 | 0.33 [0.23, 0.44] |
| Cento, f196.1 | 139 | 0.91 [0.85, 0.96] |

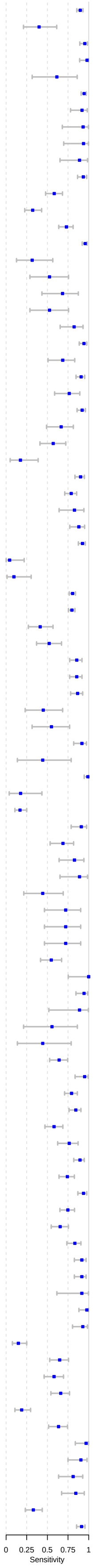

### Fig F – Forest plots for Ct-values greater 30

| Author, Study ID | Sample size | Sensitivity [95%CI] |
|---|---|---|
| Alemany, a02.ov | 103 | 0.57 [0.47, 0.67] |
| Bulilete, a07.1 | 14 | 0.14 [0.02, 0.43] |
| Gupta, a13.1 | 4 | 0.25 [0.01, 0.81] |
| Merino−Amador, a25.1 | 41 | 0.51 [0.35, 0.67] |
| Nalumansi, a27.1 | 44 | 0.54 [0.39, 0.70] |
| Nalumansi, a27.1 | 9 | 0.56 [0.21, 0.86] |
| Takeda, a50.1 | 12 | 0.17 [0.02, 0.48] |
| Krüger, a52.ov | 12 | 0.33 [0.10, 0.65] |
| Favresse, f31.1 | 16 | 0.19 [0.04, 0.46] |
| Favresse, f31.1 | 3 | 0.33 [0.01, 0.91] |
| Favresse, f31.2 | 3 | 0.00 [0.00, 0.71] |
| Favresse, f31.2 | 16 | 0.06 [0.00, 0.30] |
| Favresse, f31.3 | 3 | 0.67 [0.09, 0.99] |
| Favresse, f31.3 | 16 | 0.19 [0.04, 0.46] |
| Favresse, f31.4 | 3 | 0.00 [0.00, 0.71] |
| Favresse, f31.4 | 16 | 0.06 [0.00, 0.30] |
| Pena, f36.1 | 12 | 0.08 [0.00, 0.38] |
| Jääskeläinen, f50.1 | 25 | 0.12 [0.03, 0.31] |
| Jääskeläinen, f50.2 | 26 | 0.31 [0.14, 0.52] |
| Jääskeläinen, f50.3 | 26 | 0.38 [0.20, 0.59] |
| Okoye, f51.1 | 9 | 0.00 [0.00, 0.34] |
| Okoye, f51.1 | 3 | 0.00 [0.00, 0.71] |
| Young, f56.1 | 3 | 0.00 [0.00, 0.71] |
| Krüger, f58.1 | 24 | 0.42 [0.22, 0.63] |
| Bouassa, f67.1 | 23 | 0.70 [0.47, 0.87] |
| Sberna, f83.1 | 21 | 0.00 [0.00, 0.16] |
| Thakur, f88.1 | 3 | 0.00 [0.00, 0.71] |
| Pérez...García, f111.1 | 40 | 0.05 [0.01, 0.17] |
| Pérez...García, f111.2 | 40 | 0.17 [0.07, 0.33] |
| Nordgren, f117.1 | 23 | 0.13 [0.03, 0.34] |
| Nordgren, f117.2 | 23 | 0.30 [0.13, 0.53] |
| Fernandez−Montero, f126.1 | 8 | 0.12 [0.00, 0.53] |
| García−Fiñana, f131.1 | 17 | 0.00 [0.00, 0.20] |
| Lee, f136.1 | 136 | 0.03 [0.01, 0.07] |
| Van Honacker, f143.1 | 8 | 0.50 [0.16, 0.84] |
| Van Honacker, f143.2 | 8 | 0.12 [0.00, 0.53] |
| Van Honacker, f143.3 | 8 | 0.38 [0.09, 0.76] |
| Van Honacker, f143.4 | 8 | 0.25 [0.03, 0.65] |
| Van Honacker, f143.5 | 8 | 0.38 [0.09, 0.76] |
| Van Honacker, f143.6 | 84 | 0.05 [0.01, 0.12] |
| Kurihara, f152.1 | 25 | 0.20 [0.07, 0.41] |
| Kim, f155.1 | 11 | 0.64 [0.31, 0.89] |
| Baccani, f165.2 | 14 | 0.00 [0.00, 0.23] |
| Baccani, f165.3 | 14 | 0.00 [0.00, 0.23] |
| Mayanskiy, f188.1 | 34 | 0.85 [0.69, 0.95] |
| Fourati, f190.1 | 89 | 0.01 [0.00, 0.06] |
| Fourati, f190.2 | 89 | 0.11 [0.06, 0.20] |
| Fourati, f190.3 | 89 | 0.08 [0.03, 0.16] |
| Fourati, f190.4 | 89 | 0.09 [0.04, 0.17] |
| Fourati, f190.5 | 89 | 0.01 [0.00, 0.06] |
| Fourati, f190.6 | 89 | 0.09 [0.04, 0.17] |
| Orsi, f191.1 | 5 | 0.40 [0.05, 0.85] |
| Orsi, f191.2 | 5 | 0.00 [0.00, 0.52] |
| Suzuki, f194.1 | 14 | 0.00 [0.00, 0.23] |
| Suzuki, f194.2 | 14 | 0.21 [0.05, 0.51] |
| Thirion−Romero, f195.1 | 69 | 0.20 [0.12, 0.32] |
| Cento, f196.1 | 59 | 0.34 [0.22, 0.47] |

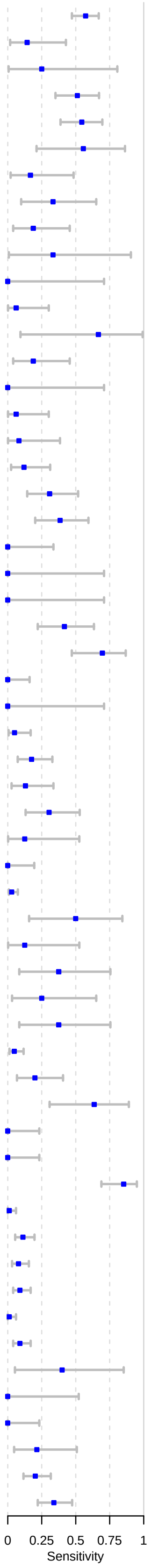
