## Supplementary material for "Accuracy of rapid point-of-care antigen-based diagnostics for SARS-CoV-2: an updated systematic review and meta-analysis with meta regression analyzing influencing factors": S3 Text

**S3a - Underlying assumptions of the meta-regression models**

The directed acyclic graph (DAG) shown in Fig 1 summarizes the assumptions for identifying the causal effect of Ct-value, symptom status, sample type, and testing procedure on test sensitivity in the first set of analyses. We assume Ct-value to directly affect test sensitivity. Furthermore, we assume time since infection to affect Ct-value and symptom status, in that symptomatic study populations have a higher chance of including patients at the beginning of the disease, where viral load is high. Similarly, we assume Ct-value and testing procedure to be affected by sample type and handling. This is due to the fact that using banked samples and diluting the samples are the main reasons for non-IFU-conforming test conduction, both also potentially reducing the samples’ viral load. In addition, non-IFU-conforming studies typically use OP or saliva samples. Since OP or saliva samples show a lower sensitivity than NP or AN / MT samples, we assume this to be connected to the samples’ viral load as well.

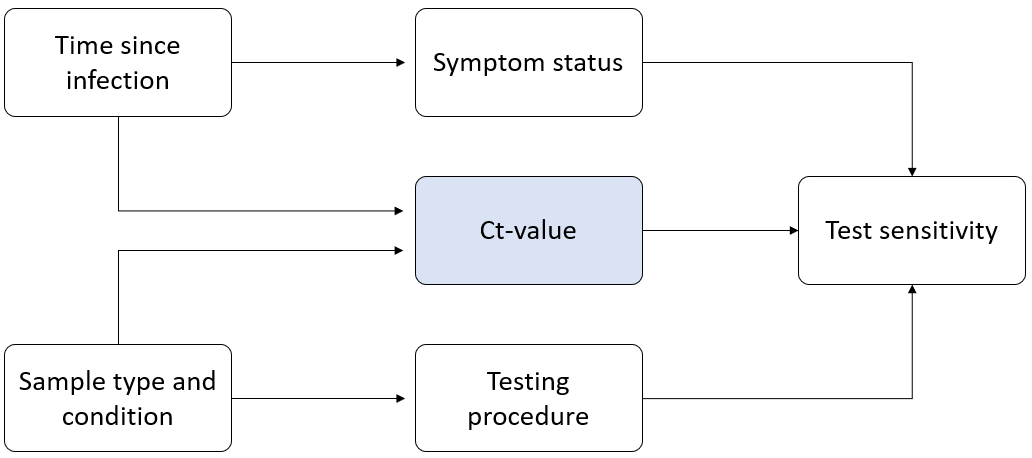

**Fig 1:** DAG for the first set of analyses

We base second and third analyses on the DAG shown in Fig 2. Due to lack of data on time since exposure for asymptomatic persons, the second and third sets of analyses are restricted to symptomatic persons. As in the first set of analyses, we assume time since infection to affect Ct-value. With time since infection being typically not reported, we consider duration of symptoms as a representative measurement. We assume the same relationships between sample type and condition, testing procedure, and Ct-value as outlined in Fig 1.

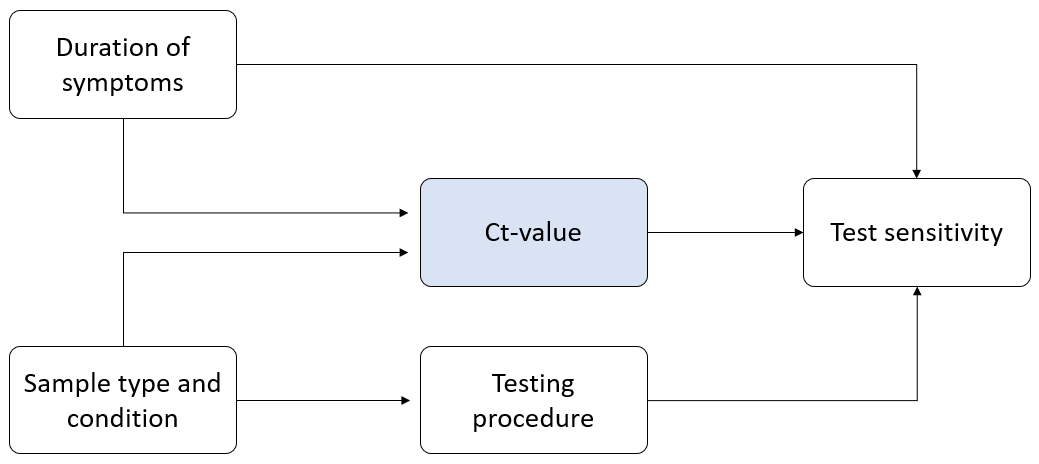

**Fig 2:** DAG for the second and third sets of analyses

**S3b - Description of Data Sets**

|  | Models estimating effects of symptom status, testing procedure, and mean Ct-value in all individuals | Models estimating effects of mean DOS, testing procedure, and mean Ct-value in symptomatic individuals | Models estimating effects of DOS > 7 days, testing procedure, and mean Ct-value in symptomatic individuals |
| --- | --- | --- | --- |
| Number of studies | 46 | 18 | 19 |
| Number of data sets | 83 | 28 | 50 |
| Number of data sets by group |  |  |  |
| Symptomatic | 65 / 83 | 28 / 28 | 50 / 50 |
| Asymptomatic | 18 / 83 | 0 / 28 | 0 / 50 |
| DOS ≤ 7 | NA | NA | 30 / 50 |
| DOS > 7 | NA | NA | 20 / 50 |
| Number of data sets by testing procedure |  |  |  |
| IFU | 54 / 83 | 21 / 28 | 40 / 50 |
| Non-IFU | 24 / 83 | 7 / 28 | 10 / 50 |
| Unclear | 5 / 83 | 0 / 28 | 0 / 50 |
| Number of data sets by sample type |  |  |  |
| AN/MT or NP | 82 / 83 | 27 / 28 | 50 / 50 |
| OP | 1 / 83 | 1 / 28 | 0 / 50 |
| Total number of observations (i.e., tests) | 10,601 | 1,627 | 2,201 |

**S3c - Additional details on the meta-regression model specification**

The unadjusted model for symptom status included an intercept and an indicator of symptom status as fixed effect terms. Data sets with the same level of study ID and symptom status shared the same value of the random effect in the unadjusted model. We used an unstructured variance-covariance matrix for the random effect corresponding to the levels of symptom status. The unadjusted models for each of the other binary variables (i.e., IFU testing procedure and DOS > 7 days) were specified in the same manner. The unadjusted models for each of the continuous variables (i.e., mean Ct-value, mean DOS) were specified in a similar manner but included random intercepts for study ID and random slopes. These random effects structures were chosen to account for between-study heterogeneity and correlations between sensitivity estimates within a study.

The adjusted models extended the unadjusted models by additionally including fixed effect terms for all confounders necessary to satisfy conditional exchangeability according to the assumed causal diagrams above. Specifically, the adjusted estimate of the effect of symptom status controlled for testing procedure and Ct-value; the adjusted estimate of the effect of testing procedure controlled for Ct-value and either symptom status (in analysis 1) or duration of symptoms (in analyses 2 and 3); the adjusted estimate of the effect of Ct-value controlled for testing procedure and either symptom status (in analysis 1) or duration of symptoms (in analyses 2 and 3); the adjusted estimate of the direct effect of duration of symptoms controlled for testing procedure and Ct-value.

**S3d – Results:** *Models estimating effects of symptom status, testing procedure, and mean Ct-value in all individuals*

Estimated Regression Coefficients:

| Effect | **Unadjusted** | **Adjusted** |
| --- | --- | --- |
| Symptomatic | 0.200 (0.137, 0.263) | 0.111 (0.048, 0.174) |
| IFU testing procedure | 0.074 (-0.011, 0.159) | 0.052 (-0.026, 0.130) |
| Average Ct-value | -0.038 (-0.048, -0.027) | -0.029 (-0.040, -0.017) |

**S3e – Results:** *Models estimating direct effect of mean DOS and effects of testing procedure and mean Ct-value in symptomatic individuals*

Estimated Regression Coefficients:

| Effect | **Unadjusted** | **Adjusted** |
| --- | --- | --- |
| Mean DOS | -0.032 (-0.079, 0.015) | 0.007 (-0.050, 0.064) |
| IFU testing procedure | -0.037 (-0.132, 0.058) | -0.080 (-0.184, 0.024) |
| Average Ct-value | -0.022 (-0.038, -0.006) | -0.028 (-0.056, 0.000) |

**S3f – Results:** *Models estimating direct effect of DOS > 7 days and effects of testing procedure and mean Ct-value in symptomatic individuals*

Estimated Regression Coefficients:

| Effect | **Unadjusted** | **Adjusted** |
| --- | --- | --- |
| DOS > 7 days | -0.229 (-0.354, -0.103) | -0.138 (-0.277, 0.001) |
| IFU testing procedure | -0.027 (-0.137, 0.083) | -0.032 (-0.142, 0.077) |
| Average Ct-value | -0.025 (-0.042, -0.008) | -0.013 (-0.033, 0.008) |
