## Supplementary material for "Accuracy of rapid point-of-care antigen-based diagnostics for SARS-CoV-2: an updated systematic review and meta-analysis with meta regression analyzing influencing factors": S4 Fig

S4 Fig. Forest plots for subgroup analysis by CT-values per test.
Caption: CI = confidence interval

### Fig A – Forest plots for Ct-value lower 20

| Test assessed | N datasets | Total Sample Size | Pooled Sensitivity (95% CI) |
|---|---|---|---|
| Panbio | 14 | 616 | 97.2% (95.3–99.2) |
| Standard Q | 15 | 833 | 98.1% (96.3–99.9) |

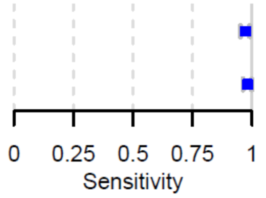


### Fig B – Forest plots for Ct-value greater 20

| Test assessed | N datasets | Total Sample Size | Pooled Sensitivity (95% CI) |
|---|---|---|---|
| Panbio | 7 | 636 | 89.2% (82.1–96.3) |
| Standard Q | 10 | 411 | 89% (81–96.9) |

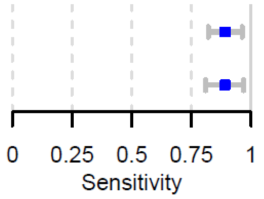


### Fig C – Forest plots for Ct-value lower 25

| Test assessed | N datasets | Total Sample Size | Pooled Sensitivity (95% CI) |
|---|---|---|---|
| Coris | 4 | 233 | 76% (58–94) |
| Innova | 4 | 214 | 75.5% (48–100) |
| Panbio | 27 | 5769 | 89.8% (85.4–94.3) |
| Standard F | 7 | 305 | 95.5% (91.3–99.8) |
| Standard Q | 21 | 2309 | 92.6% (88.5–96.7) |

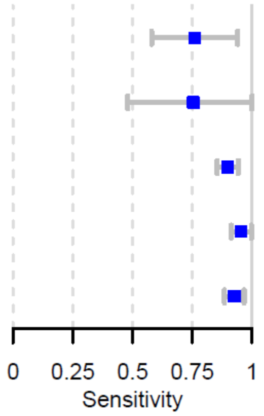


### Fig D – Forest plots for Ct-value greater 25

| Test assessed | N datasets | Total Sample Size | Pooled Sensitivity (95% CI) |
|---|---|---|---|
| Panbio | 13 | 969 | 51.2% (39.4–63) |
| Standard Q | 11 | 413 | 56.4% (45.1–67.8) |

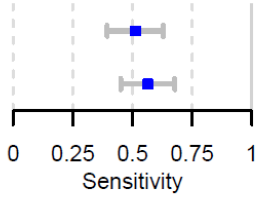


### Fig E – Forest plots for Ct-value lower 30

| Test assessed | N datasets | Total Sample Size | Pooled Sensitivity (95% CI) |
|---|---|---|---|
| BinaxNow | 6 | 332 | 83% (69.5−96.5) |
| LumiraDx | 6 | 358 | 92.6% (86.8−98.5) |
| Panbio | 31 | 5993 | 73.7% (66−81.3) |
| Standard F | 8 | 475 | 77.1% (65.8−88.3) |
| Standard Q | 25 | 5477 | 75.7% (67.9−83.4) |

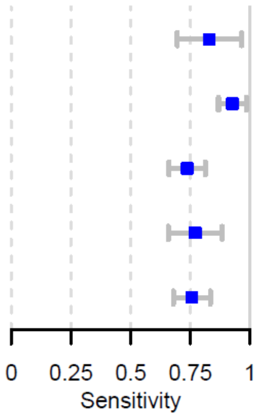


Fig F – Forest plots for Ct-value greater 30

| Test assessed | N datasets | Total Sample Size | Pooled Sensitivity (95% CI) |
|---|---|---|---|
| LumiraDx | 4 | 83 | 36% (26.1−45.9) |
| Panbio | 13 | 444 | 22.8% (12.2−33.4) |
| Standard Q | 15 | 499 | 20.4% (10.5−30.3) |

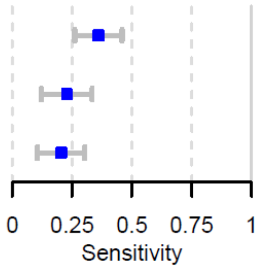
