## Supplementary material for "Accuracy of rapid point-of-care antigen-based diagnostics for SARS-CoV-2: an updated systematic review and meta-analysis with meta regression analyzing influencing factors": S4 Text

**S4 Text – Studies excluded (number of studies: 263)**

Ag-RDT not commercially available (47)

Analytical study (13)

Did not assess an Ag-RDT (25)

Duplication of data (4)

1.	Ikeda M, Imai K, Tabata S, Miyoshi K, Mizuno T, Murahara N, et al. Clinical evaluation of self-collected saliva by RT-qPCR, direct RT-qPCR, RT-LAMP, and a rapid antigen test to diagnose COVID-19. Journal of Clinical Microbiology, 2020; 59(8):e01438-20. DOI:10.1101/2020.06.06.20124123.

2.	Lindner A, Krüger L, Nikolai O, Klein JAF, Rössig H, Schnitzler P, et al. SARS-CoV-2 variant of concern B.1.1.7: diagnostic accuracy of three antigen-detecting rapid tests. medRxiv [Preprint]; published June 15, 2021. DOI:10.1101/2021.06.15.21258502.

3.	Masiá M, Fernández-González M, Sánchez M, Carvajal M, García JA, Gonzalo N, et al. Nasopharyngeal Panbio COVID-19 antigen performed at point-of-care has a high sensitivity in symptomatic and asymptomatic patients with higher risk for transmission and older age. medRxiv [Preprint]; published November 17, 2020. DOI:10.1101/2020.11.16.20230003.

4.	Klein JAF, Krüger LJ, Tobian F, Gaeddert M, Lainati F, Schnitzler P, et al. Head-to-head performance comparison of self-collected nasal versus professional-collected nasopharyngeal swab for a WHO-listed SARS-CoV-2 antigen-detecting rapid diagnostic test. Medical Microbiology and Immunology, 2021; 210(4):181-186. DOI:10.1007/s00430-021-00710-9.

Guidelines, review and modelling (50)

Monitoring (41)

No estimates for sensitivity and specificity (31)

10.1016/j.jiac.2021.06.019. Epub 2021 Jun 25.

No point of care (14)

Study population smaller 10 (3)

1.	Gandolfo Cd, Morecchiato Fd, Pistello Mp, Rossolini GMp, Cusi MGp. Detection of SARS-CoV-2 N protein allelic variants by rapid high-throughput CLEIA antigen assay. Journal of Clinical Virology, 2021; 142:104942. DOI:10.1016/j.jcv.2021.104942.

2.	Itoh K, Kawamitsu T, Osaka Y, Sato K, Suzuki Y, Kiriba C, et al. False positive results in severe acute respiratory coronavirus 2 (SARS-CoV-2) rapid antigen tests for inpatients. Journal of Infection and Chemotherapy, 2021; 27(7):1089-1091. DOI:10.1016/j.jiac.2021.03.011.

3.	Kashiwagi K, Ishii Y, Aoki K, Yagi S, Maeda T, Miyazaki T, et al. Immunochromatographic test for the detection of SARS-CoV-2 in saliva. Journal of Infection and Chemotherapy, 2020; 27(2):384-386. DOI:10.1101/2020.05.20.20107631.

Use case not diagnosis (31)

Excluded from FIND website – duplication of data (3)

1.	Foundation for Innovative New Diagnostics. FIND Evaluation of Abbott Panbio COVID-19 Ag Rapid Test Device. External Report Version 21, 10 December, 2020.

2.	Foundation for Innovative New Diagnostics. FIND Evaluation of Coris BioConcept COVID-19 Ag Respi-Strip. External Report Version 12, 10 December, 2020.

3.	Foundation for Innovative New Diagnostics. FIND Evaluation of Shenzhen Bioeasy Biotechnology Co. Ltd. 2019-nCoV Ag Rapid Test Kit (Fluorescence). External Report Version 10, 11 February 2021, 2021.

Excluded from FIND website – monitoring (1)

1.	Foundation for Innovative New Diagnostics. FIND Evaluation of SD Biosensor, Inc.; STANDARD™ F COVID-19 Ag FIA. External Report Site Specific Report Version 10, 27 April 2021, 2021.
