## Supplementary material for "Accuracy of rapid point-of-care antigen-based diagnostics for SARS-CoV-2: an updated systematic review and meta-analysis with meta regression analyzing influencing factors": S7 Fig

### S7 Fig. Forest plots for subgroup analysis by symptomatic vs. asymptomatic.

Caption: CI = confidence interval

#### Figure A - Forest plots of asymptomatic patients

| Author, Study ID | Sample size | Sensitivity [95%CI] |
|---|---|---|
| Alemany, a02.ov | 117 | 0.80 [0.71, 0.86] |
| Bulilete, a07.1 | 53 | 0.57 [0.42, 0.70] |
| Fenollar. F., a11.1 | 22 | 0.46 [0.24, 0.68] |
| Gupta, a13.1 | 13 | 0.69 [0.39, 0.91] |
| Krüger, a17.1 | 7 | 0.57 [0.18, 0.90] |
| Linares, a20.1 | 10 | 0.50 [0.19, 0.81] |
| Merino−Amador, a25.1 | 31 | 1.00 [0.89, 1.00] |
| Scohy, a34.1 | 14 | 0.29 [0.08, 0.58] |
| Shrestha, a36.1 | 47 | 0.85 [0.72, 0.94] |
| Torres, a37.1 | 79 | 0.48 [0.37, 0.60] |
| Krüger, a52.ov | 14 | 0.86 [0.57, 0.98] |
| Agullo, a56.2 | 35 | 0.23 [0.10, 0.40] |
| Agullo, a56.3 | 32 | 0.09 [0.02, 0.25] |
| Parada−Ricart, a58.1 | 1 | 0.00 [0.00, 0.98] |
| FINDdx, a62.2 | 8 | 0.25 [0.03, 0.65] |
| FINDdx, a63.2 | 1 | 0.00 [0.00, 0.98] |
| Turcato, f09.1 | 54 | 0.50 [0.36, 0.64] |
| Takeuchi, f12.1 | 33 | 0.76 [0.58, 0.89] |
| Pollock, f17.1 | 57 | 0.70 [0.57, 0.82] |
| Pollock, f17.1 | 107 | 0.65 [0.56, 0.74] |
| Halfon, f18.1 | 35 | 0.57 [0.39, 0.74] |
| James, f23.1 | 128 | 0.52 [0.43, 0.60] |
| Torres, f29.ov | 25 | 0.60 [0.39, 0.79] |
| Baro, f33.1 | 101 | 0.39 [0.29, 0.49] |
| Baro, f33.2 | 101 | 0.52 [0.41, 0.62] |
| Baro, f33.3 | 101 | 0.44 [0.34, 0.54] |
| Baro, f33.4 | 101 | 0.46 [0.36, 0.56] |
| Baro, f33.5 | 101 | 0.29 [0.20, 0.39] |
| Pena, f36.1 | 73 | 0.70 [0.58, 0.80] |
| FINDdx, f42.1 | 8 | 0.62 [0.24, 0.92] |
| FINDdx, f42.2 | 8 | 0.88 [0.47, 1.00] |
| Okoye, f51.1 | 45 | 0.53 [0.38, 0.68] |
| Krüger, f58.1 | 9 | 0.78 [0.40, 0.97] |
| Pollock, f59.1 | 128 | 0.50 [0.41, 0.59] |
| Pollock, f59.1 | 37 | 0.51 [0.34, 0.68] |

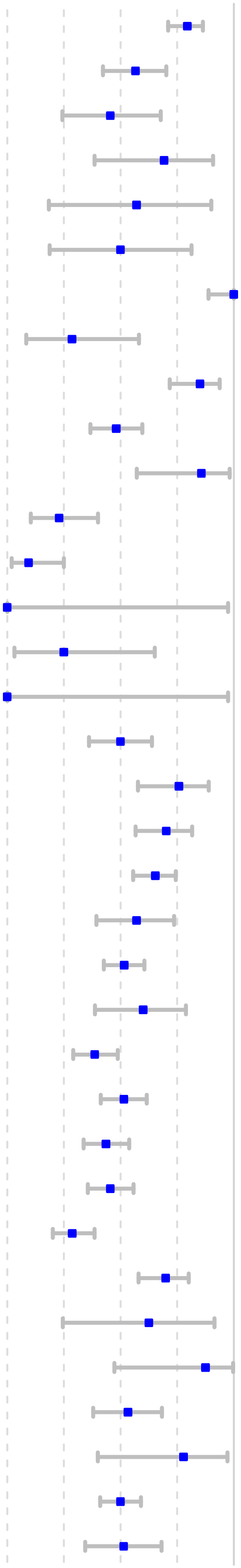

| | | |
|---|---|---|
| Stokes, f65.1 | 268 | 0.86 [0.81, 0.90] |
| Del Vecchio, f66.1 | 10 | 0.90 [0.56, 1.00] |
| L...Huillier, f72.1 | 30 | 0.43 [0.26, 0.63] |
| Osmanodja, f79.1 | 1 | 0.00 [0.00, 0.98] |
| Thakur, f88.1 | 84 | 0.34 [0.24, 0.46] |
| FINDdx, f92.1 | 15 | 0.47 [0.21, 0.73] |
| FINDdx, f93.1 | 15 | 0.73 [0.45, 0.92] |
| Brihn, f98.1 | 81 | 0.60 [0.49, 0.71] |
| Homza, f99.1 | 28 | 0.46 [0.28, 0.66] |
| Bianco, f102.1 | 101 | 0.92 [0.85, 0.96] |
| Chiu, f107.3 | 38 | 0.84 [0.69, 0.94] |
| Smith, f122.1 | 42 | 0.79 [0.63, 0.90] |
| Abdul−Mumin, f123.1 | 18 | 0.72 [0.47, 0.90] |
| Fernandez−Montero, f126.1 | 49 | 0.71 [0.57, 0.83] |
| Fernández, f128.1 | 9 | 0.78 [0.40, 0.97] |
| García−Fiñana, f131.1 | 70 | 0.40 [0.28, 0.52] |
| Shah, f133.1 | 48 | 0.69 [0.54, 0.81] |
| Di Domenico, f140.1 | 36 | 1.00 [0.90, 1.00] |
| Di Domenico, f140.2 | 36 | 0.56 [0.38, 0.72] |
| Menchinelli, f145.1 | 66 | 0.27 [0.17, 0.40] |
| Leli, f151.1 | 54 | 0.48 [0.34, 0.62] |
| Kurihara, f152.1 | 47 | 0.64 [0.48, 0.77] |
| Kim, f155.1 | 4 | 1.00 [0.40, 1.00] |
| Tinker, f156.1 | 40 | 0.20 [0.09, 0.36] |
| Merino−Amador, f160.1 | 25 | 0.92 [0.74, 0.99] |
| Allan−Blitz, f161.1 | 958 | 0.44 [0.41, 0.47] |
| Kanaujia, f162.1 | 41 | 0.56 [0.40, 0.72] |
| Kahn, f167.1 | 42 | 0.48 [0.32, 0.64] |
| Ifko, f169.1 | 16 | 0.56 [0.30, 0.80] |
| Jegerlehner, f172.1 | 25 | 0.44 [0.24, 0.65] |
| FINDdx, f180.1 | 10 | 1.00 [0.69, 1.00] |
| Kim, f186.1 | 4 | 0.75 [0.19, 0.99] |
| Thirion−Romero, f195.1 | 57 | 0.26 [0.16, 0.40] |

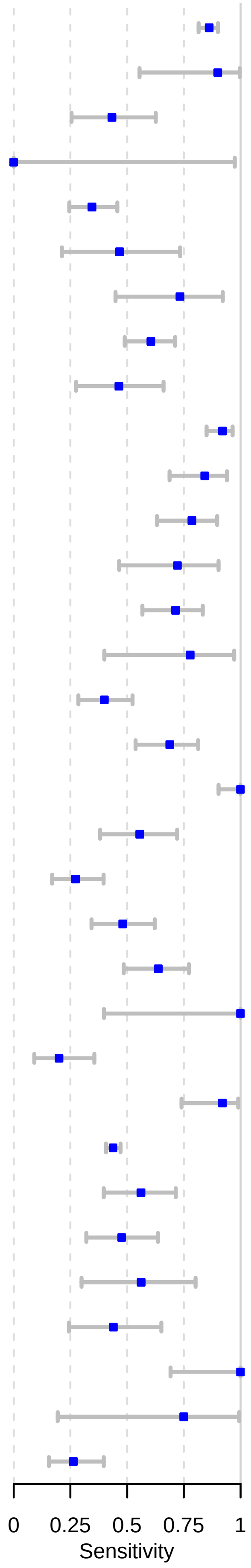

### Figure B - Forest plots of symptomatic patients

| Author, Study ID | Sample size | Sensitivity [95%CI] |
|---|---|---|
| Abdulrahman, a01.1 | 733 | 0.82 [0.79, 0.85] |
| Alemany, a02.ov | 419 | 0.93 [0.90, 0.95] |
| Albert, a03.1 | 54 | 0.80 [0.66, 0.89] |
| Beck, a04.1 | 61 | 0.77 [0.64, 0.87] |
| Bulilete, a07.1 | 87 | 0.80 [0.71, 0.88] |
| Cerutti, a08.1 | 104 | 0.72 [0.62, 0.80] |
| Fenollar. F., a11.1 | 182 | 0.79 [0.72, 0.85] |
| Gupta, a13.1 | 64 | 0.84 [0.73, 0.92] |
| Krüger, a17.1 | 40 | 0.80 [0.64, 0.91] |
| Linares, a20.1 | 50 | 0.78 [0.64, 0.88] |
| Lindner, a21.1 | 39 | 0.74 [0.58, 0.87] |
| Lindner, a21.2 | 39 | 0.80 [0.64, 0.91] |
| Merino−Amador, a25.1 | 328 | 0.90 [0.86, 0.93] |
| Pekosz, a28.1 | 28 | 0.96 [0.82, 1.00] |
| Porte, a31.1 | 82 | 0.94 [0.86, 0.98] |
| Scohy, a34.1 | 77 | 0.32 [0.22, 0.44] |
| Schwob, a35.1 | 112 | 0.93 [0.86, 0.97] |
| Schwob, a35.2 | 122 | 0.86 [0.79, 0.92] |
| Schwob, a35.3 | 138 | 0.84 [0.77, 0.90] |
| Weitzel, a41.1 | 79 | 0.62 [0.50, 0.73] |
| Weitzel, a41.3 | 78 | 0.17 [0.09, 0.27] |
| Weitzel, a41.4 | 80 | 0.85 [0.75, 0.92] |
| Young, a43.1 | 38 | 0.76 [0.60, 0.89] |
| Herrera, a46.1 | 459 | 0.77 [0.72, 0.80] |
| Mertens, a48.1 | 132 | 0.58 [0.49, 0.66] |
| Krüger, a52.ov | 92 | 0.87 [0.78, 0.93] |
| Lindner, a53.1 | 39 | 0.72 [0.55, 0.85] |
| Lindner, a53.2 | 39 | 0.80 [0.64, 0.91] |
| Agullo, a56.2 | 97 | 0.53 [0.42, 0.63] |
| Agullo, a56.3 | 89 | 0.28 [0.19, 0.39] |
| Parada−Ricart, a58.1 | 25 | 0.76 [0.55, 0.91] |
| Chaimao, a57.1 | 56 | 0.98 [0.90, 1.00] |
| FINDdx, a61.1 | 102 | 0.89 [0.81, 0.94] |
| FINDdx, a62.2 | 17 | 0.65 [0.38, 0.86] |
| FINDdx, a63.2 | 38 | 0.71 [0.54, 0.85] |
| Turcato, f09.1 | 169 | 0.90 [0.84, 0.94] |
| Takeuchi, f12.1 | 72 | 0.92 [0.83, 0.97] |
| Filgueiras, f14.1 | 54 | 0.69 [0.54, 0.80] |

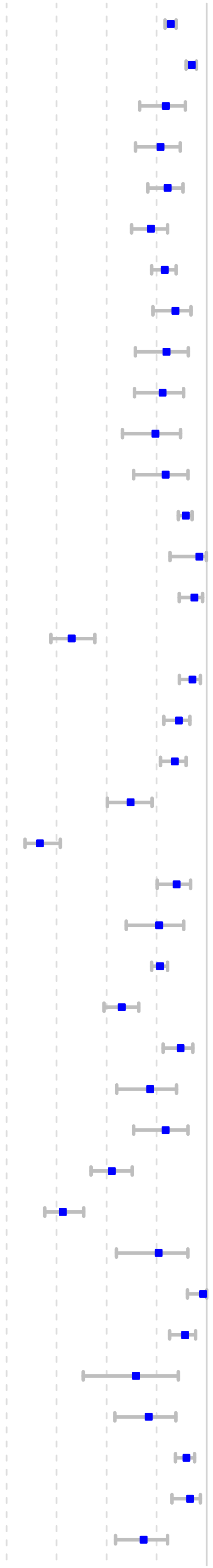

| | | | |
|---|---|---|---|
| Lindner, f15.1 | 40 | 0.85 [0.70, 0.94] | 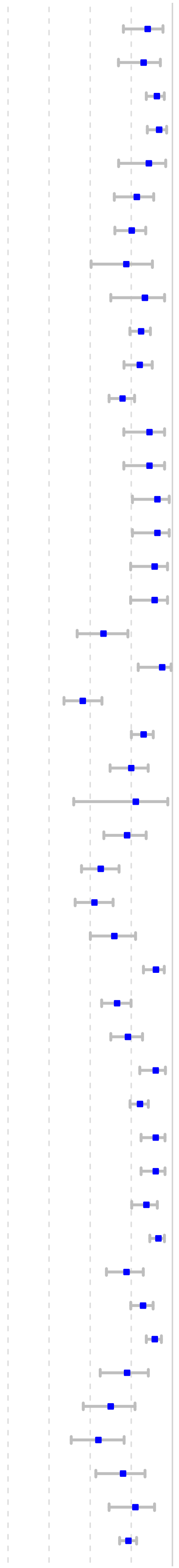 |
| Lindner, f15.2 | 40 | 0.82 [0.67, 0.93] | |
| Pollock, f17.1 | 128 | 0.91 [0.84, 0.95] | |
| Pollock, f17.1 | 100 | 0.92 [0.85, 0.96] | |
| Pollock, f17.1 | 28 | 0.86 [0.67, 0.96] | |
| Halfon, f18.1 | 51 | 0.78 [0.65, 0.89] | |
| Möckel, f19.1 | 89 | 0.75 [0.65, 0.84] | |
| Möckel, f19.2 | 25 | 0.72 [0.51, 0.88] | |
| James, f23.1 | 24 | 0.83 [0.63, 0.95] | |
| Ngo Nsoga, f28.1 | 168 | 0.81 [0.74, 0.87] | |
| Torres, f29.ov | 91 | 0.80 [0.71, 0.88] | |
| Akingba, f30.1 | 145 | 0.70 [0.62, 0.77] | |
| Nikolai, f35.1 | 36 | 0.86 [0.70, 0.95] | |
| Nikolai, f35.2 | 36 | 0.86 [0.70, 0.95] | |
| Nikolai, f35.3 | 33 | 0.91 [0.76, 0.98] | |
| Nikolai, f35.4 | 33 | 0.91 [0.76, 0.98] | |
| FINDdx, f42.1 | 37 | 0.89 [0.75, 0.97] | |
| FINDdx, f42.2 | 37 | 0.89 [0.75, 0.97] | |
| Ristic, f44.1 | 43 | 0.58 [0.42, 0.73] | |
| Marti, f46.2 | 32 | 0.94 [0.79, 0.99] | |
| Villaverde, f55.1 | 77 | 0.46 [0.34, 0.57] | |
| Krüger, f58.1 | 137 | 0.82 [0.75, 0.88] | |
| Pollock, f59.1 | 60 | 0.75 [0.62, 0.85] | |
| Pollock, f59.1 | 9 | 0.78 [0.40, 0.97] | |
| Takeuchi, f60.1 | 51 | 0.72 [0.58, 0.84] | |
| Shidlovskaya, f61.1 | 78 | 0.56 [0.45, 0.68] | |
| Shidlovskaya, f61.2 | 78 | 0.53 [0.41, 0.64] | |
| Del Vecchio, f66.1 | 51 | 0.65 [0.50, 0.78] | |
| Bouassa, f67.1 | 100 | 0.90 [0.82, 0.95] | |
| Kilic, f71.1 | 116 | 0.66 [0.57, 0.75] | |
| L...Huillier, f72.1 | 89 | 0.73 [0.63, 0.82] | |
| Osmanodja, f79.1 | 69 | 0.90 [0.80, 0.96] | |
| Thell, f81.1 | 213 | 0.80 [0.74, 0.85] | |
| FINDdx, f91.1 | 79 | 0.90 [0.81, 0.96] | |
| FINDdx, f91.2 | 79 | 0.90 [0.81, 0.96] | |
| FINDdx, f92.1 | 95 | 0.84 [0.75, 0.91] | |
| FINDdx, f93.1 | 176 | 0.92 [0.86, 0.95] | |
| Brihn, f98.1 | 68 | 0.72 [0.60, 0.82] | |
| Homza, f99.1 | 135 | 0.82 [0.75, 0.88] | |
| Bianco, f102.1 | 197 | 0.89 [0.84, 0.93] | |
| Koeleman, f103.1 | 40 | 0.72 [0.56, 0.85] | |
| Koeleman, f103.2 | 40 | 0.62 [0.46, 0.77] | |
| Koeleman, f103.3 | 40 | 0.55 [0.38, 0.71] | |
| Koeleman, f103.4 | 40 | 0.70 [0.54, 0.83] | |
| Koeleman, f103.5 | 40 | 0.78 [0.62, 0.89] | |
| Koeleman, f103.6 | 300 | 0.73 [0.68, 0.78] | |

| | | |
|---|---|---|
| Shaikh, f105.1 | 39 | 0.85 [0.70, 0.94] |
| Chiu, f107.1 | 75 | 0.85 [0.75, 0.92] |
| Chiu, f107.2 | 75 | 0.83 [0.72, 0.90] |
| Christensen, f109.1 | 60 | 0.87 [0.75, 0.94] |
| Pérez...García, f111.1 | 128 | 0.63 [0.54, 0.72] |
| Pérez...García, f111.2 | 128 | 0.68 [0.59, 0.76] |
| Harris, f114.1 | 305 | 0.82 [0.78, 0.86] |
| Nordgren, f117.1 | 156 | 0.72 [0.64, 0.79] |
| Nordgren, f117.2 | 156 | 0.80 [0.72, 0.86] |
| Smith, f122.1 | 193 | 0.76 [0.70, 0.82] |
| Abdul−Mumin, f123.1 | 24 | 0.58 [0.37, 0.78] |
| Leixner, f125.1 | 94 | 0.69 [0.59, 0.78] |
| Fernández, f128.1 | 15 | 0.93 [0.68, 1.00] |
| Terpos, f130.1 | 114 | 0.90 [0.82, 0.94] |
| Shah, f133.1 | 281 | 0.79 [0.73, 0.83] |
| Lunca, f138.1 | 40 | 0.58 [0.41, 0.73] |
| Frediani, f139.5 | 77 | 0.74 [0.63, 0.83] |
| Carbonell−Sahuquillo, f141.1 | 34 | 0.71 [0.52, 0.85] |
| Menchinelli, f145.1 | 134 | 0.89 [0.82, 0.94] |
| Cassuto, f149.1 | 32 | 0.97 [0.84, 1.00] |
| Leli, f151.1 | 112 | 0.79 [0.70, 0.86] |
| Kurihara, f152.1 | 36 | 0.89 [0.74, 0.97] |
| Bachmann, f154.1 | 108 | 0.69 [0.60, 0.78] |
| Bachmann, f154.2 | 110 | 0.74 [0.64, 0.82] |
| Bachmann, f154.3 | 109 | 0.82 [0.73, 0.88] |
| Bachmann, f154.4 | 90 | 0.83 [0.74, 0.90] |
| Bachmann, f154.5 | 92 | 0.86 [0.77, 0.92] |
| Bachmann, f154.6 | 92 | 0.91 [0.84, 0.96] |
| Kim, f155.1 | 61 | 0.88 [0.78, 0.95] |
| Allan−Blitz, f161.1 | 2107 | 0.52 [0.50, 0.54] |
| Kanaujia, f162.1 | 148 | 0.76 [0.69, 0.83] |
| Kahn, f167.1 | 51 | 0.67 [0.52, 0.79] |
| Kolwijck, f168.1 | 45 | 0.87 [0.73, 0.95] |
| Ifko, f169.1 | 23 | 0.87 [0.66, 0.97] |
| Johnson, f170.1 | 50 | 0.94 [0.84, 0.99] |
| Jegerlehner, f172.1 | 116 | 0.70 [0.61, 0.78] |
| FINDdx, f176.1 | 54 | 0.61 [0.47, 0.74] |
| FINDdx, f177.1 | 92 | 0.54 [0.44, 0.65] |
| FINDdx, f177.2 | 54 | 0.72 [0.58, 0.84] |
| FINDdx, f179.1 | 118 | 0.61 [0.52, 0.70] |
| FINDdx, f180.1 | 89 | 0.91 [0.83, 0.96] |
| FINDdx, f181.2 | 100 | 0.71 [0.61, 0.80] |
| FINDdx, f182.1 | 78 | 0.85 [0.75, 0.92] |
| FINDdx, f182.2 | 78 | 0.85 [0.75, 0.92] |
| Kim, f186.1 | 26 | 0.92 [0.75, 0.99] |
| Kim, f186.2 | 100 | 0.94 [0.87, 0.98] |
| Fourati, f190.1 | 297 | 0.35 [0.30, 0.41] |
| Fourati, f190.2 | 297 | 0.61 [0.55, 0.66] |
| Fourati, f190.3 | 297 | 0.55 [0.49, 0.61] |
| Fourati, f190.4 | 297 | 0.62 [0.56, 0.67] |
| Fourati, f190.5 | 297 | 0.33 [0.27, 0.38] |
| Fourati, f190.6 | 297 | 0.60 [0.54, 0.66] |
| Orsi, f191.1 | 60 | 0.93 [0.84, 0.98] |
| Orsi, f191.2 | 60 | 0.87 [0.75, 0.94] |
| Suzuki, f194.1 | 47 | 0.85 [0.72, 0.94] |
| Suzuki, f194.2 | 47 | 0.89 [0.77, 0.96] |
| Thirion−Romero, f195.1 | 415 | 0.58 [0.53, 0.63] |
| Koskinen, f197.1 | 13 | 1.00 [0.75, 1.00] |

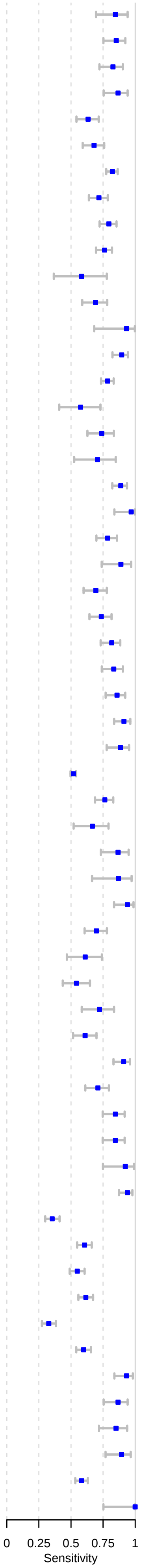

Figure C - Forest plots of patients with symptom onset greater than seven days

| Author, Study ID | Sample size | Sensitivity [95%CI] |
|---|---|---|
| Gupta, a13.1 | 7 | 0.71 [0.29, 0.96] |
| Krüger, a17.1 | 4 | 1.00 [0.40, 1.00] |
| Linares, a20.1 | 13 | 0.54 [0.25, 0.81] |
| Lindner, a21.1 | 6 | 0.17 [0.00, 0.64] |
| Lindner, a21.2 | 6 | 0.33 [0.04, 0.78] |
| Porte, a31.1 | 5 | 0.80 [0.28, 1.00] |
| Krüger, a52.ov | 13 | 0.62 [0.32, 0.86] |
| Lindner, a53.1 | 7 | 0.29 [0.04, 0.71] |
| Lindner, a53.2 | 7 | 0.71 [0.29, 0.96] |
| Chaimao, a57.1 | 12 | 1.00 [0.74, 1.00] |
| FINDdx, a62.2 | 2 | 0.50 [0.01, 0.99] |
| FINDdx, a63.2 | 6 | 0.17 [0.00, 0.64] |
| Filgueiras, f14.1 | 11 | 0.36 [0.11, 0.69] |
| Lindner, f15.1 | 5 | 0.40 [0.05, 0.85] |
| Lindner, f15.2 | 5 | 0.40 [0.05, 0.85] |
| Pollock, f17.1 | 15 | 0.67 [0.38, 0.88] |
| Pollock, f17.1 | 2 | 1.00 [0.16, 1.00] |
| Nikolai, f35.3 | 3 | 0.67 [0.09, 0.99] |
| Nikolai, f35.4 | 3 | 0.67 [0.09, 0.99] |
| FINDdx, f42.1 | 2 | 0.50 [0.01, 0.99] |
| FINDdx, f42.2 | 2 | 0.50 [0.01, 0.99] |
| Drain, f43.1 | 10 | 0.90 [0.56, 1.00] |
| Drain, f43.2 | 6 | 0.83 [0.36, 1.00] |
| Pérez−García, f52.1 | 55 | 0.49 [0.35, 0.63] |
| Pérez−García, f52.2 | 55 | 0.58 [0.44, 0.71] |
| Krüger, f58.1 | 15 | 0.47 [0.21, 0.73] |
| Pollock, f59.1 | 9 | 0.22 [0.03, 0.60] |
| Pollock, f59.1 | 2 | 0.50 [0.01, 0.99] |
| Shidlovskaya, f61.1 | 34 | 0.47 [0.30, 0.65] |
| Shidlovskaya, f61.2 | 34 | 0.41 [0.25, 0.59] |
| Del Vecchio, f66.1 | 12 | 0.33 [0.10, 0.65] |
| Thell, f81.1 | 24 | 0.62 [0.41, 0.81] |
| FINDdx, f92.1 | 6 | 0.33 [0.04, 0.78] |
| FINDdx, f93.1 | 12 | 0.75 [0.43, 0.94] |
| Pérez...García, f111.1 | 55 | 0.31 [0.19, 0.45] |
| Pérez...García, f111.2 | 55 | 0.38 [0.25, 0.52] |
| Kim, f155.1 | 20 | 0.75 [0.51, 0.91] |
| FINDdx, f180.1 | 11 | 0.54 [0.23, 0.83] |
| Fourati, f190.1 | 89 | 0.16 [0.09, 0.25] |
| Fourati, f190.2 | 89 | 0.37 [0.27, 0.48] |
| Fourati, f190.3 | 89 | 0.35 [0.25, 0.46] |
| Fourati, f190.4 | 89 | 0.42 [0.31, 0.52] |
| Fourati, f190.5 | 89 | 0.15 [0.08, 0.24] |
| Fourati, f190.6 | 89 | 0.43 [0.32, 0.54] |
| Orsi, f191.1 | 15 | 0.87 [0.60, 0.98] |
| Orsi, f191.2 | 15 | 0.80 [0.52, 0.96] |
| Thirion−Romero, f195.1 | 139 | 0.37 [0.29, 0.45] |

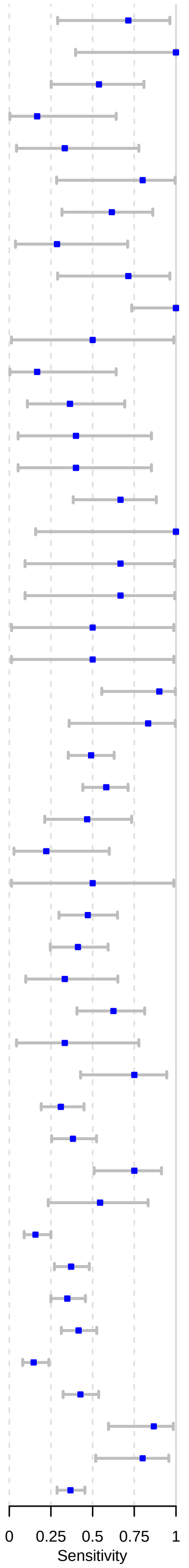

Figure D - Forest plots of patients with symptom onset less than seven days

| Author, Study ID | Sample size | Sensitivity [95%CI] |
|---|---|---|
| Gupta, a13.1 | 57 | 0.86 [0.74, 0.94] |
| Krüger, a17.1 | 34 | 0.79 [0.62, 0.91] |
| Linares, a20.1 | 37 | 0.86 [0.71, 0.96] |
| Lindner, a21.1 | 33 | 0.85 [0.68, 0.95] |
| Lindner, a21.2 | 33 | 0.88 [0.72, 0.97] |
| Pekosz, a28.1 | 28 | 0.96 [0.82, 1.00] |
| Porte, a31.1 | 76 | 0.95 [0.87, 0.98] |
| Young, a43.1 | 38 | 0.76 [0.60, 0.89] |
| Krüger, a52.ov | 76 | 0.91 [0.82, 0.96] |
| Lindner, a53.1 | 32 | 0.81 [0.64, 0.93] |
| Lindner, a53.2 | 32 | 0.81 [0.64, 0.93] |
| Chaimao, a57.1 | 44 | 0.98 [0.88, 1.00] |
| FINDdx, a62.2 | 13 | 0.77 [0.46, 0.95] |
| FINDdx, a63.2 | 32 | 0.81 [0.64, 0.93] |
| Filgueiras, f14.1 | 43 | 0.77 [0.61, 0.88] |
| Lindner, f15.1 | 35 | 0.89 [0.73, 0.97] |
| Lindner, f15.2 | 35 | 0.91 [0.77, 0.98] |
| Pollock, f17.1 | 85 | 0.96 [0.90, 0.99] |
| Pollock, f17.1 | 26 | 0.85 [0.65, 0.96] |
| Nikolai, f35.1 | 36 | 0.86 [0.70, 0.95] |
| Nikolai, f35.2 | 36 | 0.86 [0.70, 0.95] |
| Nikolai, f35.3 | 30 | 0.93 [0.78, 0.99] |
| Nikolai, f35.4 | 30 | 0.93 [0.78, 0.99] |
| FINDdx, f42.1 | 34 | 0.94 [0.80, 0.99] |
| FINDdx, f42.2 | 34 | 0.91 [0.76, 0.98] |
| Drain, f43.1 | 73 | 0.99 [0.93, 1.00] |
| Drain, f43.2 | 34 | 1.00 [0.90, 1.00] |
| Pérez−García, f52.1 | 46 | 0.85 [0.71, 0.94] |
| Pérez−García, f52.2 | 46 | 0.91 [0.79, 0.98] |
| Villaverde, f55.1 | 77 | 0.46 [0.34, 0.57] |
| Krüger, f58.1 | 118 | 0.86 [0.79, 0.92] |
| Pollock, f59.1 | 51 | 0.84 [0.71, 0.93] |
| Pollock, f59.1 | 7 | 0.86 [0.42, 1.00] |
| Shidlovskaya, f61.1 | 44 | 0.64 [0.48, 0.78] |
| Shidlovskaya, f61.2 | 44 | 0.61 [0.46, 0.76] |
| Del Vecchio, f66.1 | 39 | 0.74 [0.58, 0.87] |
| Osmanodja, f79.1 | 69 | 0.90 [0.80, 0.96] |
| Thell, f81.1 | 189 | 0.82 [0.76, 0.88] |

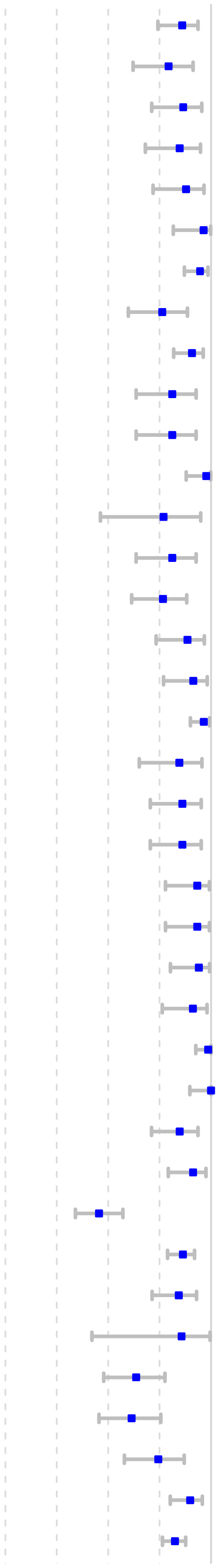

| | | |
|---|---|---|
| FINDdx, f92.1 | 87 | 0.88 [0.80, 0.94] |
| FINDdx, f93.1 | 162 | 0.93 [0.88, 0.97] |
| Pérez...García, f111.1 | 73 | 0.88 [0.78, 0.94] |
| Pérez...García, f111.2 | 73 | 0.90 [0.81, 0.96] |
| Terpos, f130.1 | 114 | 0.90 [0.82, 0.94] |
| Shah, f133.1 | 243 | 0.82 [0.76, 0.86] |
| Frediani, f139.5 | 77 | 0.74 [0.63, 0.83] |
| Carbonell−Sahuquillo, f141.1 | 34 | 0.71 [0.52, 0.85] |
| Bachmann, f154.1 | 108 | 0.69 [0.60, 0.78] |
| Bachmann, f154.2 | 110 | 0.74 [0.64, 0.82] |
| Bachmann, f154.3 | 109 | 0.82 [0.73, 0.88] |
| Bachmann, f154.4 | 90 | 0.83 [0.74, 0.90] |
| Bachmann, f154.5 | 92 | 0.86 [0.77, 0.92] |
| Bachmann, f154.6 | 92 | 0.91 [0.84, 0.96] |
| Kim, f155.1 | 41 | 0.95 [0.84, 0.99] |
| FINDdx, f175.1 | 51 | 0.92 [0.81, 0.98] |
| FINDdx, f176.1 | 50 | 0.62 [0.47, 0.75] |
| FINDdx, f177.1 | 85 | 0.54 [0.43, 0.65] |
| FINDdx, f177.2 | 45 | 0.78 [0.63, 0.89] |
| FINDdx, f178.1 | 101 | 0.90 [0.82, 0.95] |
| FINDdx, f179.1 | 111 | 0.61 [0.52, 0.70] |
| FINDdx, f180.1 | 78 | 0.96 [0.89, 0.99] |
| FINDdx, f180.2 | 29 | 0.76 [0.56, 0.90] |
| FINDdx, f181.1 | 96 | 0.58 [0.48, 0.68] |
| FINDdx, f181.2 | 70 | 0.81 [0.70, 0.90] |
| FINDdx, f182.1 | 66 | 0.82 [0.70, 0.90] |
| FINDdx, f182.2 | 66 | 0.82 [0.70, 0.90] |
| Fourati, f190.1 | 200 | 0.45 [0.38, 0.52] |
| Fourati, f190.2 | 200 | 0.71 [0.64, 0.77] |
| Fourati, f190.3 | 200 | 0.66 [0.58, 0.72] |
| Fourati, f190.4 | 200 | 0.71 [0.64, 0.77] |
| Fourati, f190.5 | 200 | 0.41 [0.34, 0.48] |
| Fourati, f190.6 | 200 | 0.69 [0.62, 0.75] |
| Orsi, f191.1 | 45 | 0.96 [0.85, 1.00] |
| Orsi, f191.2 | 45 | 0.89 [0.76, 0.96] |
| Thirion−Romero, f195.1 | 276 | 0.70 [0.64, 0.75] |

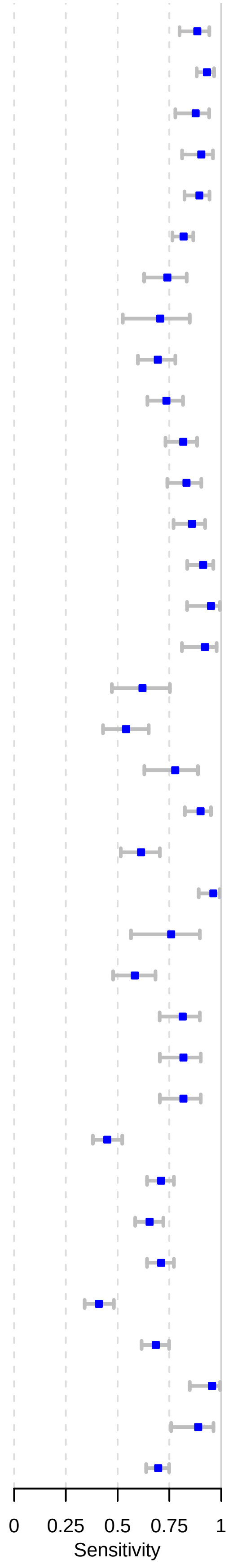
