## Supplementary material for "Accuracy of rapid point-of-care antigen-based diagnostics for SARS-CoV-2: an updated systematic review and meta-analysis with meta regression analyzing influencing factors": S8 Fig

S8 Fig. Forest plot for subgroup analysis by mean Ct-values for TP and FN samples

Caption: CI = confidence interval

| Subgroup | N Datasets | Total Sample Size | Pooled Mean Ct−value (95% CI) |
|---|---|---|---|
| True Positives | 69 | 5083 | 22.2 (21.5, 22.8) |
| False Negatives | 78 | 2408 | 30.4 (29.7, 31.1) |

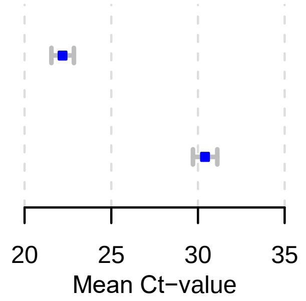
