## Supplementary material for "Accuracy of rapid point-of-care antigen-based diagnostics for SARS-CoV-2: an updated systematic review and meta-analysis with meta regression analyzing influencing factors": S9 Fig

### S9 Fig. Forest plots for subgroup analysis by mean Ct-values for TP and FN samples

Caption: CI = confidence interval

#### Figure A – Forest plots of mean Ct-value for true positive samples

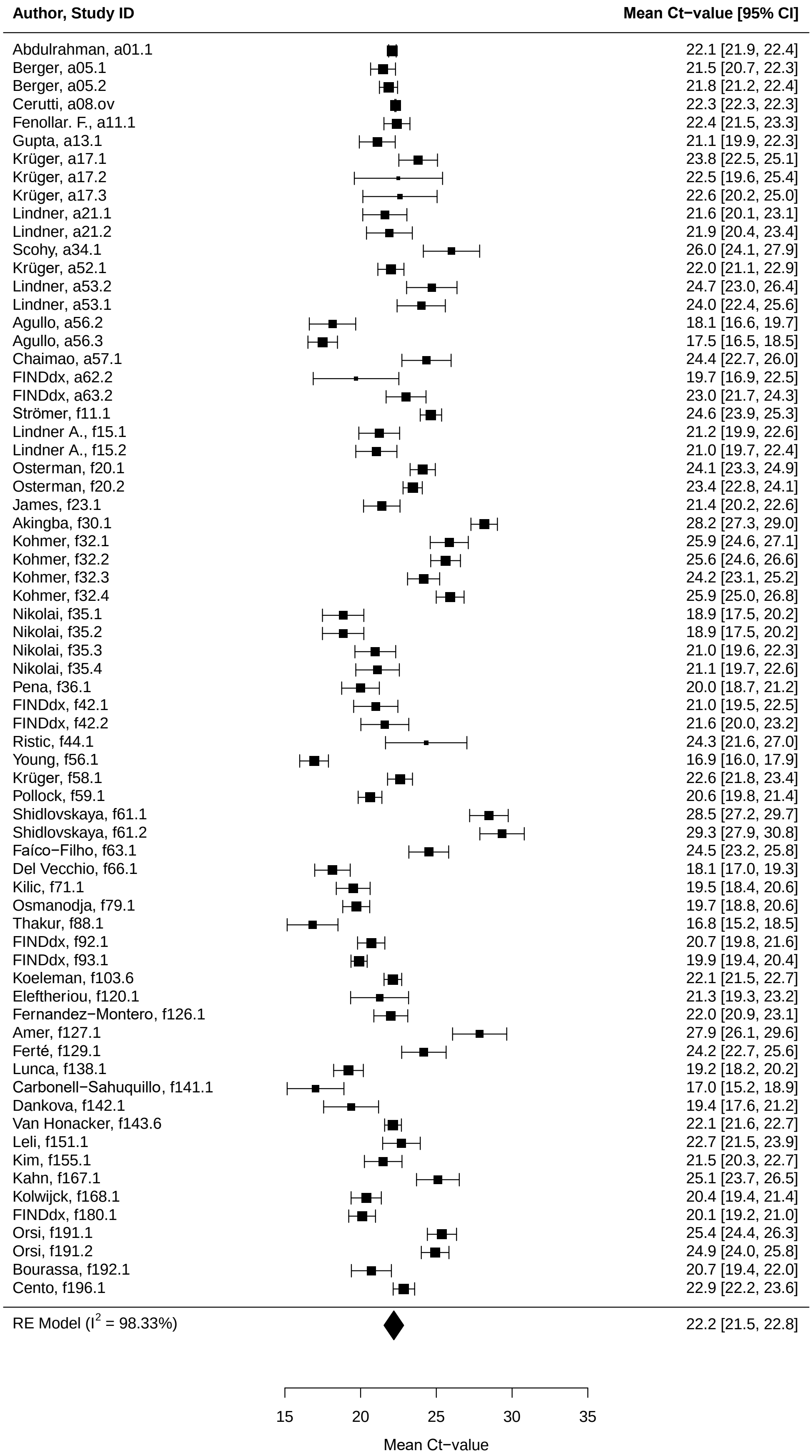

Figure B – Forest plots of mean Ct-value for false negative samples

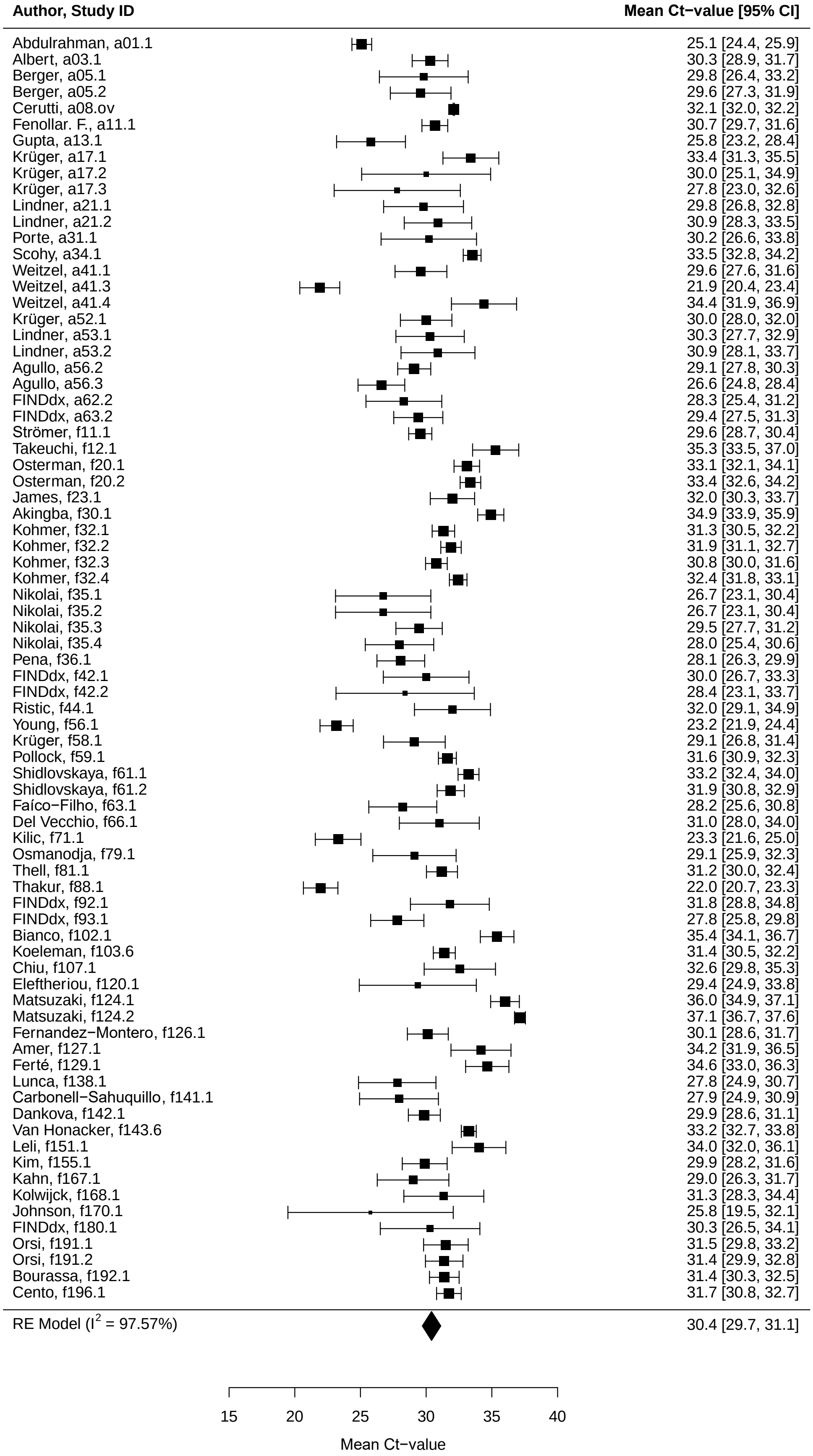
