## Supplementary material for "Accuracy of rapid point-of-care antigen-based diagnostics for SARS-CoV-2: an updated systematic review and meta-analysis with meta regression analyzing influencing factors": S10 Fig

S10 Fig. HSROC curve Standard Q nasal and LumiraDx Ag-RDT

Caption: HSROC = Hierarchical summary receiver-operating characteristic

Fig A – HSROC curve Standard Q nasal

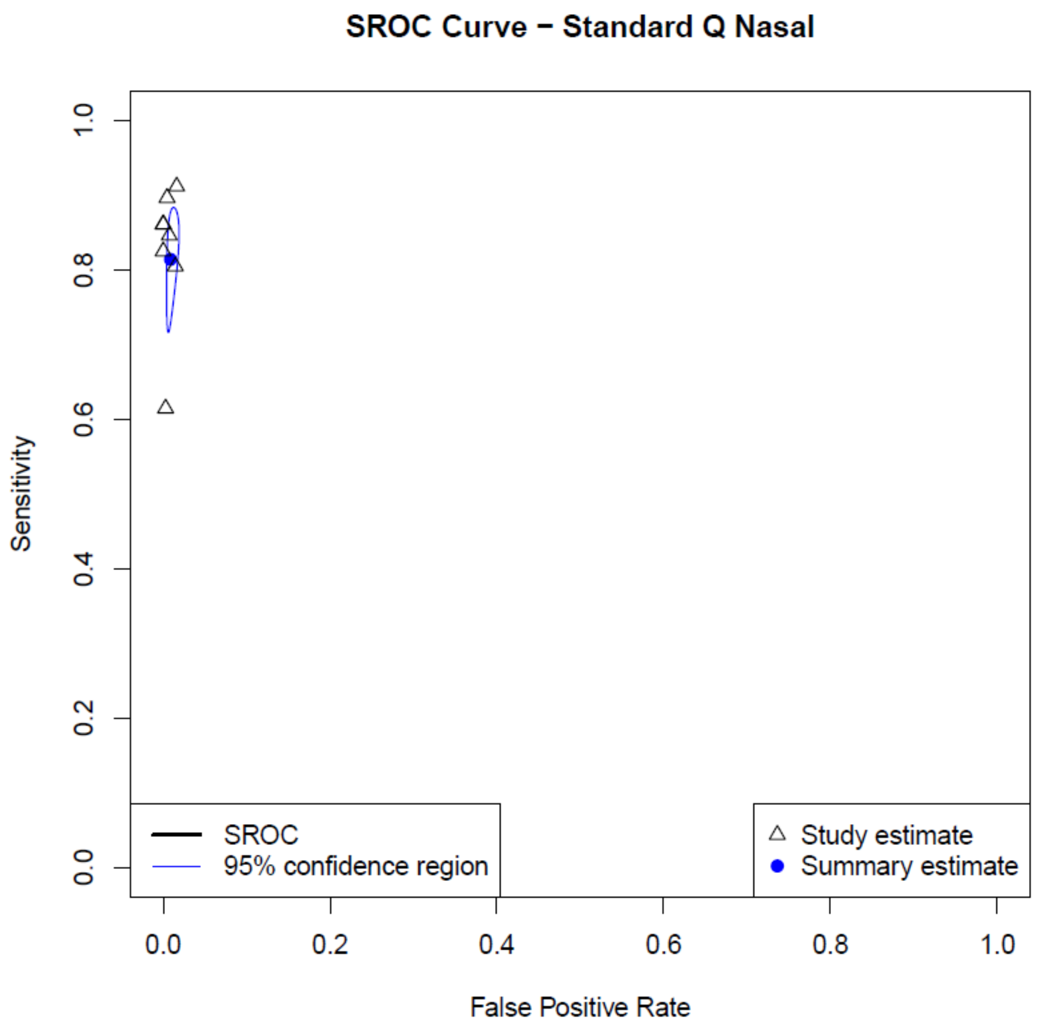

Fig B – HSROC curve LumiraDx

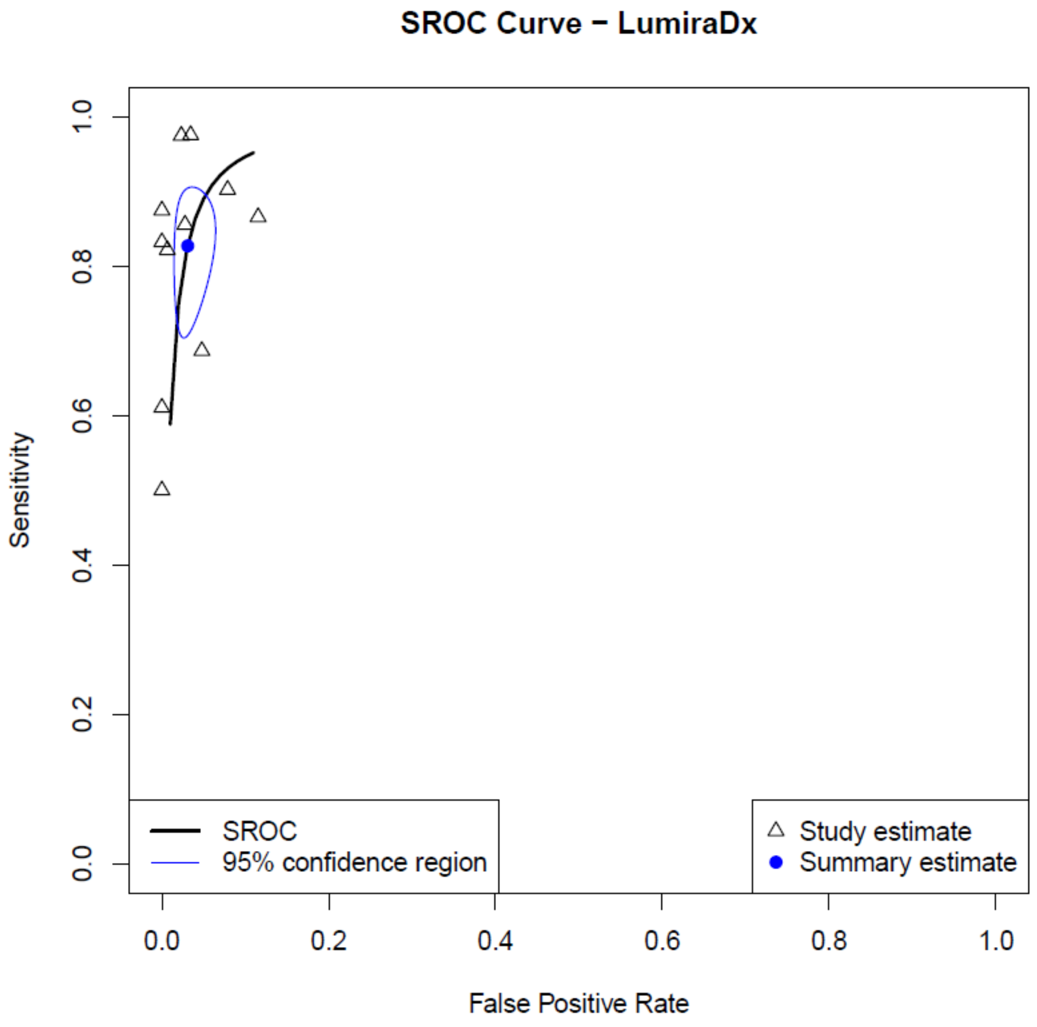
