## Supplementary material for "Accuracy of rapid point-of-care antigen-based diagnostics for SARS-CoV-2: an updated systematic review and meta-analysis with meta regression analyzing influencing factors": S11 Fig

S11 Fig. Forest plot for univariate analysis for Nadal and SureScreen V

Caption: CI = confidence interval

| Test assessed | Sensitivity N datasets | Sensitivity Total Sample Size | Pooled Sensitivity (95% CI) | Specificity N datasets | Specificity Total Sample Size | Pooled Specificity (95% CI) |
|---|---|---|---|---|---|---|
| Nadal | 5 | 1026 | 56.8% (29.6−84) | 4 | 902 | 91.1% (80.2−100) |
| SureScreen V | 5 | 555 | 58% (38.3−77.6) | | | |

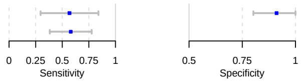
